# Collaborative nowcasting of COVID-19 hospitalization incidences in Germany

**DOI:** 10.1101/2023.04.17.23288668

**Authors:** Daniel Wolffram, Sam Abbott, Matthias an der Heiden, Sebastian Funk, Felix Günther, Davide Hailer, Stefan Heyder, Thomas Hotz, Jan van de Kassteele, Helmut Küchenhoff, Sören Müller-Hansen, Diellë Syliqi, Alexander Ullrich, Maximilian Weigert, Melanie Schienle, Johannes Bracher

## Abstract

Real-time surveillance is a crucial element in the response to infectious disease outbreaks. However, the interpretation of incidence data is often hampered by delays occurring at various stages of data gathering and reporting. As a result, recent values are biased downward, which obscures current trends. Statistical nowcasting techniques can be employed to correct these biases, allowing for accurate characterization of recent developments and thus enhancing situational awareness. In this paper, we present a preregistered real-time assessment of eight nowcasting approaches, applied by independent research teams to German 7-day hospitalization incidences. This indicator played an important role in the management of the pandemic in Germany and was linked to levels of non-pharmaceutical interventions via certain thresholds. Due to its definition, in which hospitalization counts are aggregated by the date of case report rather than admission, German hospitalization incidences are particularly affected by delays and can take several weeks or months to fully stabilize. For this study, all methods were applied from 22 November 2021 to 29 April 2022, with probabilistic nowcasts produced each day for the current and 28 preceding days. Nowcasts at the national, state, and age-group levels were collected in the form of quantiles in a public repository and displayed in a dashboard. Moreover, a mean and a median ensemble nowcast were generated. We find that overall, the compared methods were able to remove a large part of the biases introduced by delays. Most participating teams underestimated the importance of very long delays, though, resulting in nowcasts with a slight downward bias. The accompanying uncertainty intervals were also too narrow for almost all methods. Averaged over all nowcast horizons, the best performance was achieved by a model using case incidences as a covariate and taking into account longer delays than the other approaches. For the most recent days, which are often considered the most relevant in practice, a mean ensemble of the submitted nowcasts performed best. We conclude by providing some lessons learned on the definition of nowcasting targets and practical challenges.

## 1 Introduction

During infectious disease outbreaks, real-time surveillance data contributes to situational awareness and risk management, informing resource planning and control measures. However, the timely interpretation of epidemiological indicators is often hampered by the preliminary nature of real-time data. Due to reporting delays, the most recent data points are usually incomplete and subject to retrospective upward corrections.

This bias can lead to incorrect conclusions about current trends. Statistical *nowcasting* methods aim to remedy this problem by predicting how strongly preliminary data points are still going to be corrected upwards, taking into account the associated uncertainty. Nowcasts thus help to uncover current trends which are not yet visible in reported numbers.

Problems of this type have been extensively researched across various disciplines; e.g., in econometrics, the gross domestic product and the inflation rate are routinely nowcasted [1]. Methods for preliminary count data as encountered in the present work originated in the actuarial sciences, where they were developed to handle insurance claims data [2]. In epidemiology, the problem of delayed reporting has been treated in diverse contexts, including the HIV pandemic [3], foodborne Escherichia coli outbreaks [4], the 2009 influenza pandemic [5] and mosquito-borne diseases like malaria [6] and dengue [7], [8]. During the COVID-19 pandemic, the problem has seen growing interest, and new approaches tailored to a variety of settings have been suggested [9]–[14]. There is thus an ever-growing number of methods to statistically correct reporting delays. However, two important aspects are rarely addressed in the current literature. Firstly, few studies assess the performance of methods in real-time settings. The papers we are aware of – with [14] as an exception – contain only retrospective case studies which risk smoothing over some of the difficulties occurring in real time (e.g., major data revisions, time pressure on analysts). Also, few studies include comparisons with existing methods. While occasionally one additional model is applied for comparison [8], [11], [13], systematic comparative assessments are lacking. Our work fills this gap by examining multiple procedures in real time, thus providing a realistic picture of nowcast performance and the arising practical challenges. By bringing together several different models, our study is moreover the first able to assess the potential of combined ensemble nowcasts.

We evaluate the different nowcasting approaches in an application to German 7-day hospitalization incidences. These have played an important role in the management of the pandemic in Germany. Indeed, in November 2021, they were defined as the key indicator to determine levels of non-pharmaceutical interventions. Via a system of thresholds [15], they played an important role in the management of the pandemic, in particular in the fall and winter of 2021. Nowcasting is of particular importance for this indicator due to the way it was defined. As will be detailed in Section 2.1, the official German hospitalization numbers published by Robert Koch Institute (RKI) are aggregated by the reporting date of the associated positive test rather than the date of hospital admission. The total time span between the case report and the hospitalization report (i.e. the “delay” that has to be predicted) thus consists of two parts: the time between the report of the positive test and hospital admission and the actual reporting delay between hospitalization and the reporting thereof. This definition led to some criticism in the public discourse but was defended as a necessary compromise between timeliness and data quality by RKI [16]. Figure 1 illustrates the nowcasting task in the context of the 7-day hospitalization incidence. It shows real-time nowcasts from 1 December 2021, 1 February 2022, and 1 April 2022. Comparison with a more stable data version from 8 August 2022 shows that in these instances, the nowcasts were able to correctly reveal the actual trends, which differed sharply from the apparent declines found in the data as available at the time of nowcasting.

**Figure 1:**
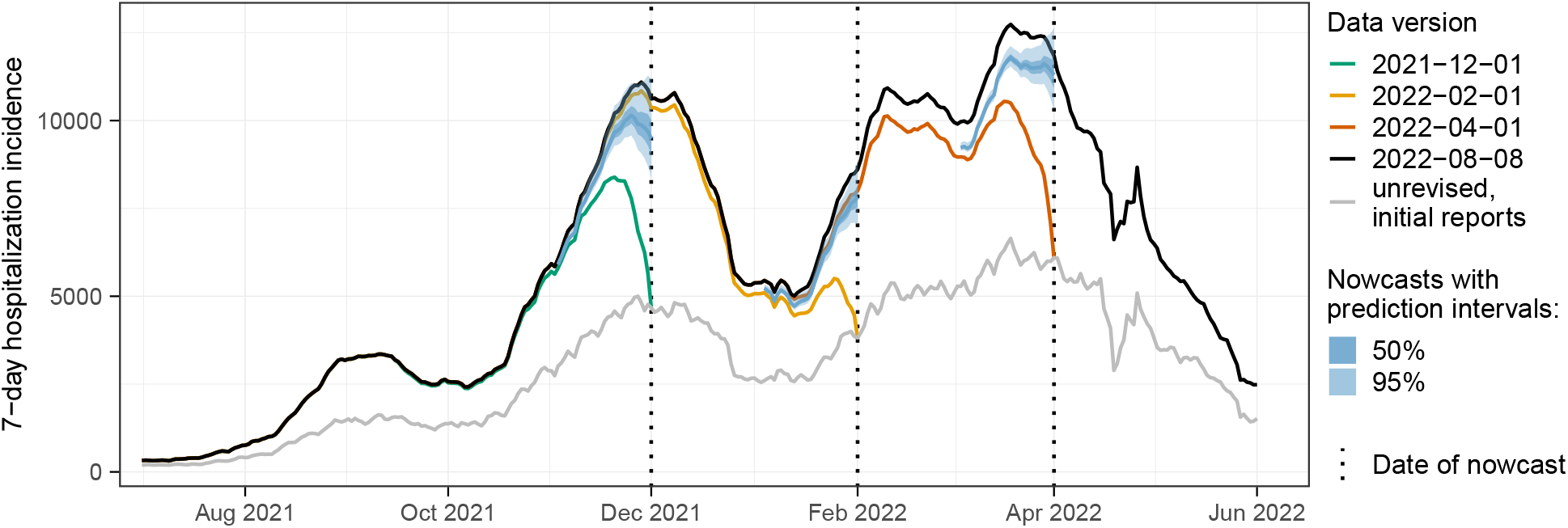
Illustration of the nowcasting task. Data as available in real time (colored lines) are incomplete and especially for recent dates considerably lower than the final corrected values (black line). Nowcasts (blue-shaded areas) aim to predict in real time what the final data points are going to be. The light grey line shows the initially reported value as available on the respective date.

The present work is based on a collaborative platform, the *COVID-19 Nowcast Hub*, which we launched soon after the hospitalization incidence became the guideline value for the German pandemic policy. It served to collect and combine real-time nowcasts from several models on a daily basis. The approach builds upon the *COVID-19 Forecast Hubs*, which during the pandemic were run in the US [17], Germany and Poland [18], and later the entire European Economic Area, Switzerland and the UK [19]. These Hubs showed that combining different epidemiological models into an ensemble can produce more robust predictions, confirming results from forecasting challenges like *FluSight* on seasonal influenza [20]. We aimed for compatibility with the Forecast Hub ecosystem in many technical and methodological aspects, in particular by following the same submission format and evaluation criteria [21]. This way we contribute to a growing evidence base on predictive epidemic modeling in real time.

The remainder of the manuscript is structured as follows. In Section 2, we introduce the 7-day hospitalization incidence as defined by RKI and outline the agreed-upon nowcast targets. We present the individual nowcasting methods and ensemble approaches, as well as the prespecified evaluation criteria. Section 3 presents the results of our formal performance evaluation, followed by qualitative observations on periods of unusual reporting patterns or the emergence of a new variant. We then assess the impact of model revisions as well as the sensitivity of the results to the exact definition of the nowcast target. Section 4 concludes with a discussion.

## 2 Methods

To facilitate a transparent assessment, we preregistered our evaluation study, specifying the criteria to assess the submitted nowcasts. The study protocol was deposited at the registry of the Open Science Foundation on 23 November 2021 [22]. In some instances, we had to deviate from the protocol. These are detailed in the respective subsections and summarized in Supplementary Table 2.

### 2.1 Definition of the COVID-19 7-day hospitalization incidence

Data on the German COVID-19 hospitalization incidence was published in a daily rhythm by Robert Koch Institute [23]. By its official definition [24], it is given by the number of hospitalized COVID-19 cases among cases reported over a 7-day period relative to 100,000 inhabitants. Hospitalizations are thus aggregated by the case reporting date, more precisely, when a case was digitally registered by a local health authority, rather than the date of hospital admission (though the two may coincide). We will refer to this case reporting date as the *reference date* in the following. We note that the hospitalization is not required to occur during the 7-day window mentioned previously, nor is COVID-19 required to be the main reason for hospitalization. When new hospitalizations are added to the record, they may thus change the value of the 7-day hospitalization incidence for past periods, depending on how much time has passed between the positive test, the time of hospitalization, and ultimately its reporting. The initially reported value of the hospitalization incidence is thus only an approximate and typically too low value.

To illustrate the extent of these revisions, Figure 2 shows the fraction of the 7-day hospitalization incidence that was reported 0 – 70 days after the respective reference date. Same-day values covered 50–60% of the ultimately reported hospitalizations, with a slight upward trend over the study period (left panel). Around 85% were reached after 14 days and even after 70 days, there were upward corrections of more than 3%. As illustrated in the second panel, same-day reporting completeness varied considerably across states. In Bremen (HB) it exceeded 75%, whereas it was below 50% in Saxony (SN) and Hamburg (HH). Reporting completeness was also variable across age groups and weekdays (third and fourth panels). A detailed display of temporal variations in initial reporting completeness across states can be found in Supplementary Figure 13. It should be noted that initial reporting completeness can also depend on the overall strain on the health system, and delays tend to be longer in times of high caseloads [25].

**Figure 2:**
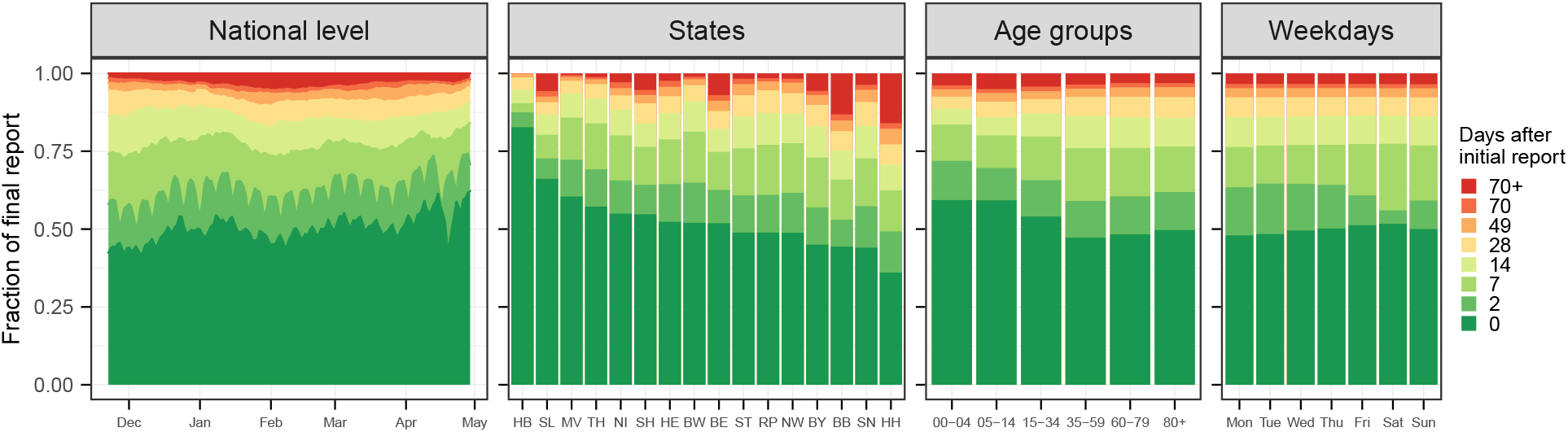
Completeness of 7-day hospitalization incidences 0 to 70 days after the respective reference date. First panel: temporal development over the considered study period, aggregated over states and age groups. Second: by state, ordered by initial reporting completeness (see Supplementary Figure 13 for the definition of abbreviations). Third: by age group. Fourth: by weekday.

As mentioned before, thresholds of 3, 6, and 9 per 100,000 population were introduced in the fall of 2021 and used to determine the necessary extent of non-pharmaceutical interventions [15]. These were applied to the initial value of the hospitalization incidence as reported on the respective day without any retrospective completion. This value is also referred to as the *frozen value*. For illustration, these frozen values were added to Figure 1 (light grey line). We note that due to the temporal and geographic differences shown in Figure 2, the same frozen value can translate to rather different final values of the hospitalization incidence.

### 2.2 Nowcast targets and study period

The goal of the collected nowcasts was to predict how much the preliminary values of the hospitalization incidence were still going to change. Specifically, on each day during the period from Monday 22 November 2021 to Friday 29 April 2022, a prediction needed to be issued for the final value the 7-day hospitalization incidence would take for that day and the previous 28 days. In the study protocol, we defined the final state to be predicted as the time series available on 8 August 2022. This date was chosen to be 100 days after the end of our study period. Originally, teams were asked to provide nowcasts for all working days of the study period, excluding a Christmas break. However, as all teams fully automated their approaches, we were able to collect nowcasts on weekends and public holidays and include them in the study.

Teams were asked to issue nowcasts for the national level as well as for the 16 German states and seven different age groups (as available in public RKI data; 00-04, 05-14, 15-34, 35-59, 60-79 and 80+ years). No age-specific nowcasts at the state level were generated. To quantify prediction uncertainty, a probabilistic format was adopted, where teams had to submit seven quantiles (2.5%, 10%, 25%, 50%, 75%, 90%, 97.5%) of the predictive distribution in addition to the mean. Following the procedure in the various COVID-19 Forecast Hubs, our main analysis treated all outcomes on their original count scales, i.e. not standardized by population. This means that the relative size of states or age strata is reflected in the weight they receive in the overall evaluation [21].

In the study protocol, we also defined a retrospective study period reaching from 1 July 2021 to 19 November 2021. The motivation was to compare the retrospective performance on historical data available during model development to prospective performance under real-world conditions. However, due to time constraints, only two teams provided complete sets of retrospective nowcasts prior to the beginning of the prospective study. We therefore chose to omit this aspect. Instead, we added an evaluation of retrospective nowcasts from four revised models to the main study period from 22 November 2021 to 29 April 2022.

### 2.3 Overview of models

Nowcasts from eight independently run models were collected for the duration of our study. Six of them were contributed by groups of academics, one by the Robert Koch Institute (`RKI`) and one by the data science team of the newspaper *Süddeutsche Zeitung* (`SZ`). A short description of the different methods is provided in Table 1. Most approaches took preliminary hospitalization numbers as their only input, applying various techniques to model delay distributions and the underlying time series of hospitalizations. Only the `ILM` model took a different approach, in which the number of confirmed cases was included as an explanatory variable. Approaches also differed in terms of the methods used for inference, uncertainty quantification, the flexibility and complexity of their delay distribution and time series models, as well as the maximum delay considered (ranging from 35 to 84 days). Some models obtained nowcasts at a coarser spatial or age resolution by hierarchically aggregating nowcasts generated for finer strata. Models were typically not fitted to the entire available data set, but only a recent subset, the size of which again differed by team.

**Table 1:** Description of nowcast models contributed to the collaborative project.

| Abbreviation | Short description | Reference | Generation of prediction intervals | Data input | Weekday effects | Max. delay | Length of training data | Hierarchical aggregation |
| --- | --- | --- | --- | --- | --- | --- | --- | --- |
| **Epiforecasts** | Bayesian model assuming the underlying curve of hospitalizations follows a random walk on the log scale. Reporting delays are assumed to follow a log-normal distribution with time-varying parameters. Report date effects are handled via a random effect for day of the week. | [26] | Bayesian posterior distribution | Hospitalizations | Yes | 40 days | 40 days | No |
| **ILM** | Hospitalization probabilities given a positive test are estimated separately per delay time and age group. These are used to predict yet unreported hospitalizations based on case incidences. | In prep. | Based on past nowcast errors | Hospitalizations, cases | Indirectly via aggregation | 84 days | 91 days | Yes |
| **KIT** | A simple multiplication factor approach, with uncertainty intervals generated by comparing past point nowcasts to observations. | Append. E | Based on past nowcast errors | Hospitalizations | No | 40 days | 60 days | No |
| **LMU** | Nowcasts are based on a generalized additive model, with the delay distribution described by a sequential multinomial model. | [27], [28] | Parametric bootstrap approach using the covariance of the estimated model parameters and a Poisson observation model | Hospitalizations | Yes | 40 days | 56 days | Partially |
| **RIVM** | Counts per reference date and delay are modeled by a two-dimensional P-spline surface and covariates including weekday effects. This surface is extrapolated to fill in yet unknown values. | [29] | Parametric bootstrap approach using the covariance of the estimated model parameters and a negative binomial observation model | Hospitalizations | Yes | 42 days | 84 days | No |
| **RKI** | The conditional reporting delay probabilities are modeled using a logistic regression model that incorporates weekday, federal state, and two age groups as covariates ($\leq 60$, $> 60$). | [30], [31] | Sampling from model-based distribution | Hospitalizations | Yes | 40 days | 68 days | Yes |
| **SU** | Similarly to **Epiforecasts**, the latent curve of daily hospitalizations is assumed to follow a random walk on the log scale. The delay distribution is modeled via a discrete-time hazard model with weekday effects for the reporting day. Entries of the reporting triangle are assumed to follow a negative binomial distribution. | [9] | Bayesian posterior distribution | Hospitalizations | Yes | 35 days | 56 days | No |
| **SZ** | Nowcasts are based on the empirical distribution of the relative difference between initially reported and retrospectively completed values of the hospitalization incidence. | – | Empirical quantiles of past relative corrections | Hospitalizations | No | – | 60 days | No |

**Table 2:** Deviations from the study protocol.

| Description | Reasoning |
| --- | --- |
| Inclusion of weekends into the study period. | Initially, weekends were excluded from the study period as we expected submission to require some human intervention at least initially. As all teams completely automated their procedures very quickly, we were able to update models on weekends and include them in the evaluation. |
| Inclusion of the Christmas period into the study period. | In the study protocol, the Christmas period was excluded from the study period as (i) we expected irregular reporting behavior and (ii) we did not want to oblige modelers to perform any manual steps for submission during this period. However, as there were no unusual patterns and all submissions were completely automated, we decided to include this period. We note that, contrary to our expectations, the Easter period showed some unusual reporting patterns. We nonetheless kept it in the evaluation as removing periods for which nowcasts were expected to work normally but did not would unduly embellish our results. |
| Omission of the retrospective study period from 1 July 2021 to 19 November 2021. | We initially planned to include a retrospective study period in order to contrast the performance of methods in retrospect and in real time. However, as only two teams provided retrospective nowcast before the start of the study period, we chose to omit this aspect. Instead, we chose to include an analysis of four revised versions of contributed models applied retrospectively to the period 22 November to 29 April. |
| Omission of nowcast targets for which even including fill-in nowcasts results could not be obtained from all methods. | For a very small set of targets we were unable to obtain submissions from all models, even including fill-in nowcasts. As these represented a negligible fraction of all targets (0.3%) we pragmatically chose to omit these from the main analysis in order to achieve a balanced data set. |
| Omission of interval coverage results at the 80% level. | As interval coverage results at the 80% level provided no additional insights and led to overly full figures we chose to omit them. |
| Definition of naïve baseline model. | At the time of writing the protocol we had decided that a naïve baseline model should be included, but it was unclear how it should be defined. The **FrozenBaseline** used in the main analysis was only defined during the work on the manuscript. |
| Tightening of ensemble inclusion criteria. | During the study period, we realized that one model (SZ) occasionally issued nowcast values below the already known values (which in almost all cases only get corrected upwards). This is due to a smoothing step that is included in the procedure. We decided to exclude these from the ensemble. Specifically, submissions were excluded from the ensemble whenever the median or mean nowcast was below the already known value. |

**Table 3:** Missingness of real-time submissions by the participating teams. Apart from the targets listed in Table 4, all of these could be imputed with fill-in nowcasts.

| Model | dates without submission |
| --- | --- |
| **Epiforecasts** | 25–26 Jan 2022 |
| ILM | 27–28 Nov 2021, 24, 26, 30 Dec 2021, 16 Jan 2022, 8–20 April 2022 |
| RIVM | 8 Dec 2021, 23 Apr 2022 |
| RKI | 8 Dec 2021; all 0 and -1 day nowcasts |
| SU | 27–28 Nov 2021, 5 Dec 2022 |

**Table 4:** Nowcast targets for which no complete sets of submissions could be obtained. These amount to 394 nowcast targets among the 109,968 considered in total.

| Dates | Excluded targets | Reason |
| --- | --- | --- |
| 22 November 2021 | Horizons -1 and 0 days, all strata | On the first day of our study several models provided only nowcasts from $-2$ days backward. |
| 22–24 November 2021 | Horizons -23 to -28 days, all strata. | The **SZ** model initially only provided nowcasts three weeks back. |
| 31 Jan, 1 Feb 2022 | State of Hamburg, all horizons. | Nowcasts from **Epinowcasts** model not available due to convergence issues. |

### 2.4 Ensemble approaches

On a daily basis, all submissions that were available at 2pm were combined to generate an ensemble nowcast, see Figure 3 for an illustration. We created the two following ensembles.

**Figure 3:**
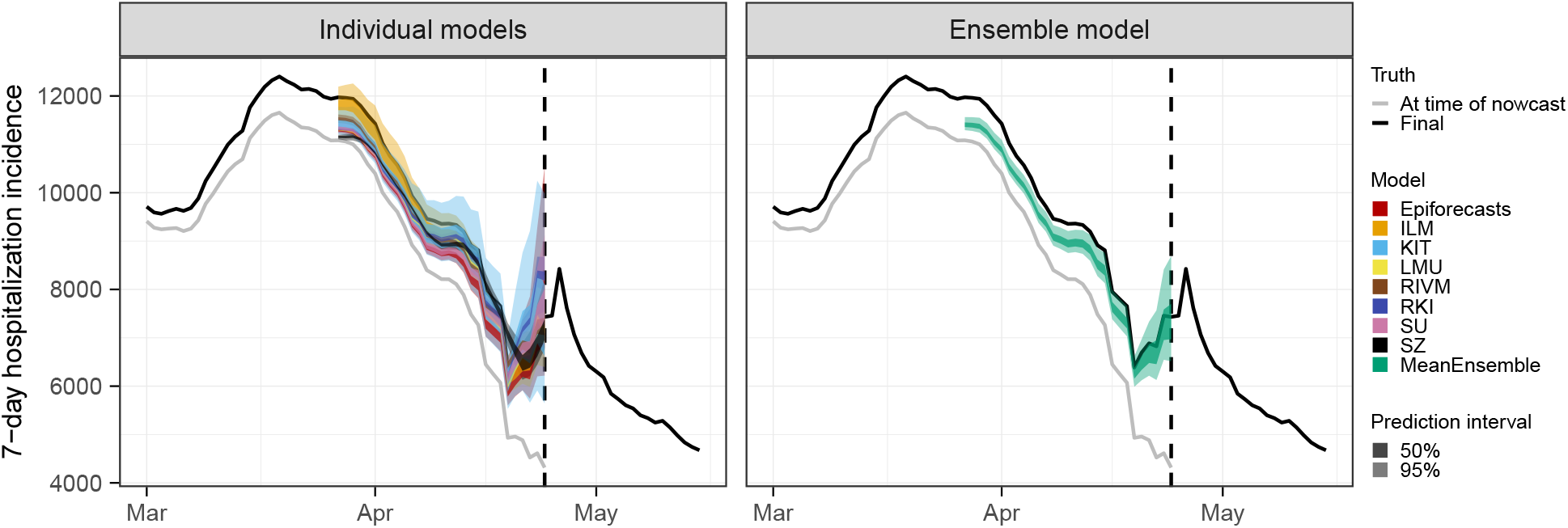
Illustration of an aggregation approach to combine a set of individual nowcasts into an ensemble nowcast. Here, the ensemble is computed as the quantile-wise mean of all submissions.

- For the `MeanEnsemble` each predictive quantile was obtained as the arithmetic mean of the respective quantiles of the member nowcasts. The ensemble mean was obtained as the mean of the submitted predictive means.
- For the `MedianEnsemble` the same procedure was applied using the median rather than the arithmetic mean for aggregation.

This direct aggregation at the level of quantiles rather than, e.g., probability density functions, is known as *Vincentization* [32]. A discussion of its properties and differences to the aggregation of density functions can be found in [33]. As the expected number of contributed models was moderate, the `MeanEnsemble` was expected to be better-behaved than the `MedianEnsemble`, which can produce oddly shaped distributions in such settings [18]. The `MeanEnsemble` was therefore prespecified as the primary ensemble approach (unlike in e.g. [17] or [18]).

### 2.5 Evaluation metrics

*Proper scoring rules* are an established tool to evaluate probabilistic forecasts [34], or, in our setting, now-casts. They are constructed such that they encourage honest forecasting, i.e., forecasters optimize their expected score by reporting their true beliefs about the future. Put differently, there is no way of “gaming” the system and obtaining improved scores by reporting modified versions of one’s actual prediction. As in our setting nowcasts consist of three nested central prediction intervals, a natural choice is the interval score [34]. For an interval [*l, u*] at the level (1 − *α*), *α* ∈ (0, 1), reaching from the 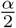- to the 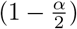-quantile of the predictive distribution *F*, it is defined as

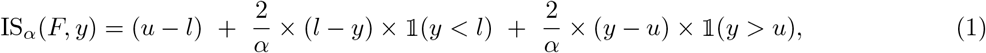

where 𝟙 is the indicator function and *y* is the realized value. Here, the first term characterizes the spread of the predictive distribution, the second penalizes overprediction (observations fall below the prediction interval) and the third term penalizes underprediction. To assess all submitted quantiles of the predictive distribution jointly we use the weighted interval score (WIS; [21]), which is a weighted average of interval scores at different nominal levels and the absolute error. For *N* nested prediction intervals it is defined as

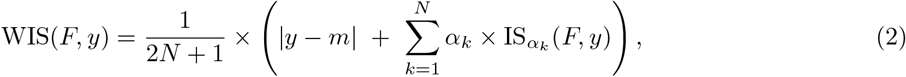

where *m* is the predictive median and in our setting *N* = 3 and *α*_1_ = 0.5, *α*_2_ = 0.2, *α*_3_ = 0.05. We note that it is equivalent to the mean pinball loss across the respective quantile levels, which is often employed in quantile regression [21]. The WIS approximates the widely used continuous ranked probability score (CRPS) and can be interpreted as a generalization of the absolute error to probabilistic predictions. It is negatively oriented, meaning that lower values are better. The decomposition of the interval score into spread, overprediction, and underprediction also translates to the WIS and can be used to enhance the interpretability of results.

To put results into perspective, we defined the simplistic baseline model `FrozenBaseline` which applies no correction and just issues the current data version as its deterministic nowcast (i.e., with all quantiles set to the same value). This allowed us to compute relative scores

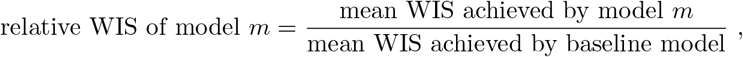

characterizing the improvement over the uncorrected time series. Here, lower values are better, and values below 1 imply that the nowcasts reduce the error of the uncorrected time series. We note that while the study protocol specified that a baseline model was to be included, its definition was only agreed upon later. We note that the `KIT` model was originally conceived as a baseline model, but later considered too complex for this purpose; in the preregistration, it is therefore referred to as a “reference model”.

To assess the central tendency of nowcasts we used the mean absolute error for predictive medians and the mean squared error for predictive means (i.e., for each functional we use the respective *consistent scoring function* [35]). To evaluate calibration, i.e., the statistical consistency between nowcasts and observations, we consider the empirical coverage of the 50% and 95% prediction intervals,

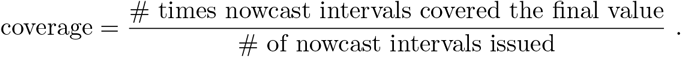

In case of missing submissions, i.e., if a team failed to provide a nowcast on a given day, nowcasts could be filled in retrospectively. To assess whether this had a substantial impact on the comparative evaluation, we applied a pairwise comparison scheme as described in [17] to compare models using only the sets of nowcast tasks treated in real time by each model. Details can be found in Supplementary Section C.

## 3 Results

### 3.1 Completeness of submissions

All participating teams produced nowcasts over the entire study period and only rarely failed to submit nowcasts in time (see Supplementary Table 4). In most cases, missing nowcasts were filled in retrospectively. In very few cases (0.3% of all targets; see Table 4) it was not possible to obtain submissions from all teams; to handle these cases we chose to slightly deviate from the study protocol and omit the respective targets in our evaluation. The `ILM` model did not provide state-level nowcasts, while the `RKI` model did not include age-stratified results. Moreover, the `RKI` model only provided point nowcasts and two quantiles in real time (at levels 2.5% and 97.5%); the remaining quantiles were only provided in retrospect. We encountered some more minor difficulties, e.g., due to missing quantiles for certain targets; we summarize these and the chosen solutions in Supplementary Table 4.

**Table 5:** Other decisions in response to unexpected difficulties.

| Description | Reasoning |
| --- | --- |
| Imputation of 0.1-quantiles from **Epiforecasts** model via an interpolation/normal approximation. | For several months the **Epiforecasts** model did not provide 0.1 quantiles, which was only noticed towards the end of the study period. In order to be able to evaluate the WIS without having to rerun the model for all concerned dates, we imputed the 0.1-quantile by interpolating between the 0.025 and the 0.25 quantile based on a normal approximation (which implies that the 0.1 quantile is almost exactly halfway between the 0.025 and 0.25 quantiles). |
| LMU nowcasts for Saarland and Bremen were replaced by nowcasts from the retrospectively revised model version discussed in Section 3.6. | In their real-time submissions, the **LMU** team only reported point nowcasts for the states of Saarland and Bremen, which are considerably smaller than the other states (1M and 700k inhabitants, respectively). To be able to nonetheless evaluate the WIS and keep these two states in the overall evaluation, we used the revised nowcasts as discussed in Section 3.6 as these contained all quantiles for these states. We consider this defensible as the role of these two states in the overall evaluation under WIS is very small (the WIS typically scales with the order of magnitude of the target). |
| Filling in some missing entries of **ILM** with nowcasts from the updated model. | Nowcasts from 22–26 and 29 November 2021 were missing entries for horizons -28, -1, and 0 days. To fill these in, we used the revised nowcasts as discussed in Section 3.6. |
| Removal of a small number of obviously erroneous nowcasts for the **RKI** model. | In a handful of instances, the **RKI** model submitted obviously erroneous nowcasts for the 0-day horizon. These stated values of more than 1 billion hospitalizations. We replaced these with the respective -1 day nowcasts. |

### 3.2 Visual inspection of nowcasts

For a first impression of nowcast performance, Figure 4 shows same-day nowcasts at the national level (i.e., at each date the respective nowcast with a horizon of 0 days is shown). Figure 5 shows the same for nowcasts 14 days back in time (i.e., for each day the nowcast issued 14 days later is shown). Displayed are the median predictions along with the central 50% and 95% prediction intervals. The light grey line shows the data as available when the nowcast was issued (which in Figure 4 corresponds to the *frozen values*), and the red line shows the respective final value as available on 8 August 2022. In both figures, it can be seen that nowcasts from all models are generally close to the final values to be predicted. However, considerable variability in interval widths is apparent, ranging from rather wide (`KIT`) to very narrow intervals (`LMU`, `RKI`). Some models, in particular `KIT` and `SZ`, display pronounced weekday patterns in their same-day nowcasts, which to a lower degree also carry through to the ensemble nowcasts. For the nowcasts 14 days back in time we observe a slight downward bias in the central tendency, the only exception being the `ILM` model. As most of the concerned models moreover feature quite narrow prediction intervals, these often do not cover the final values.

**Figure 4:**
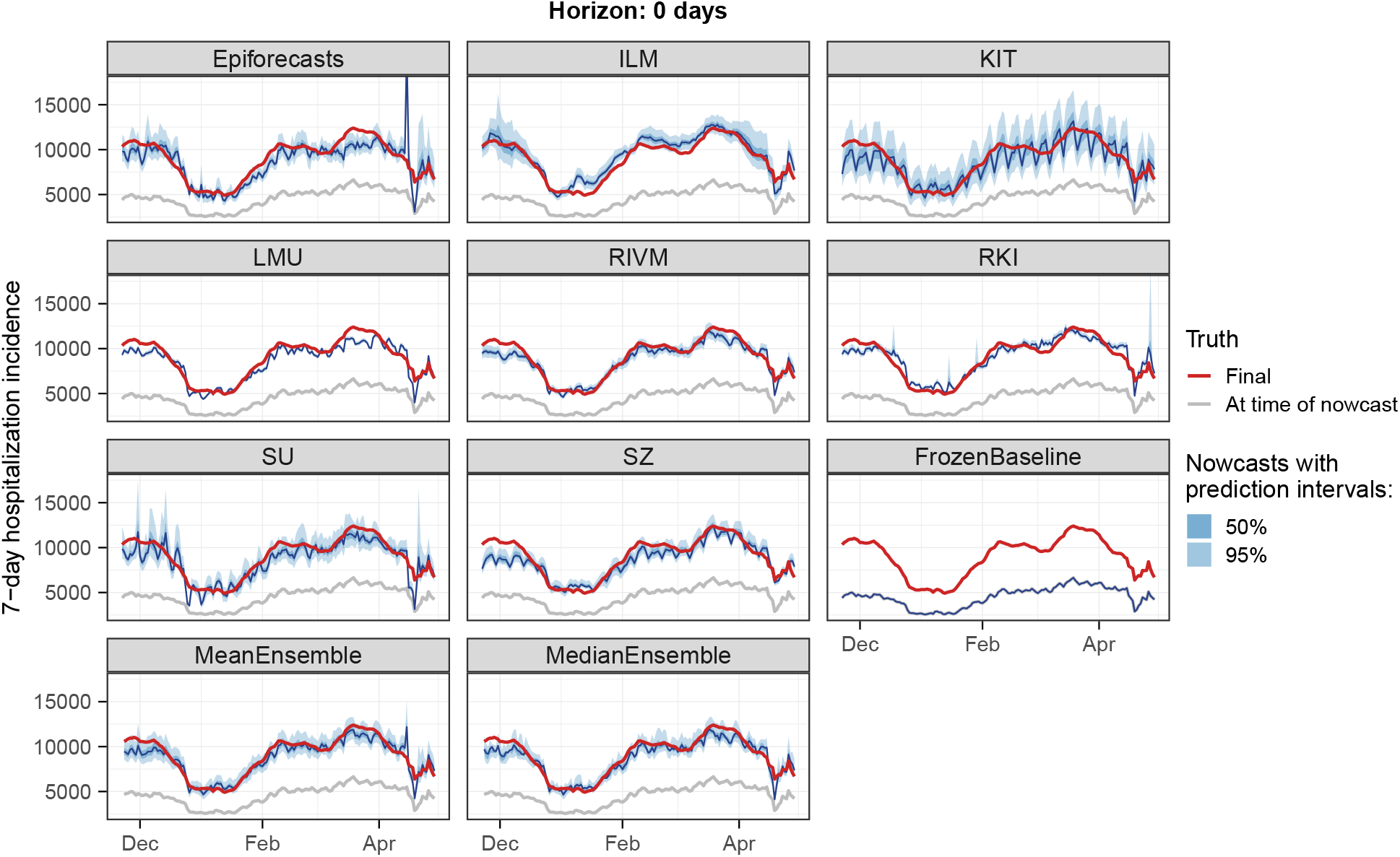
Same-day nowcasts (with a horizon of 0 days) of the 7-day hospitalization incidence as issued on each day of the study period. Nowcasts are shown for the German national level pooled across all age groups.

**Figure 5:**
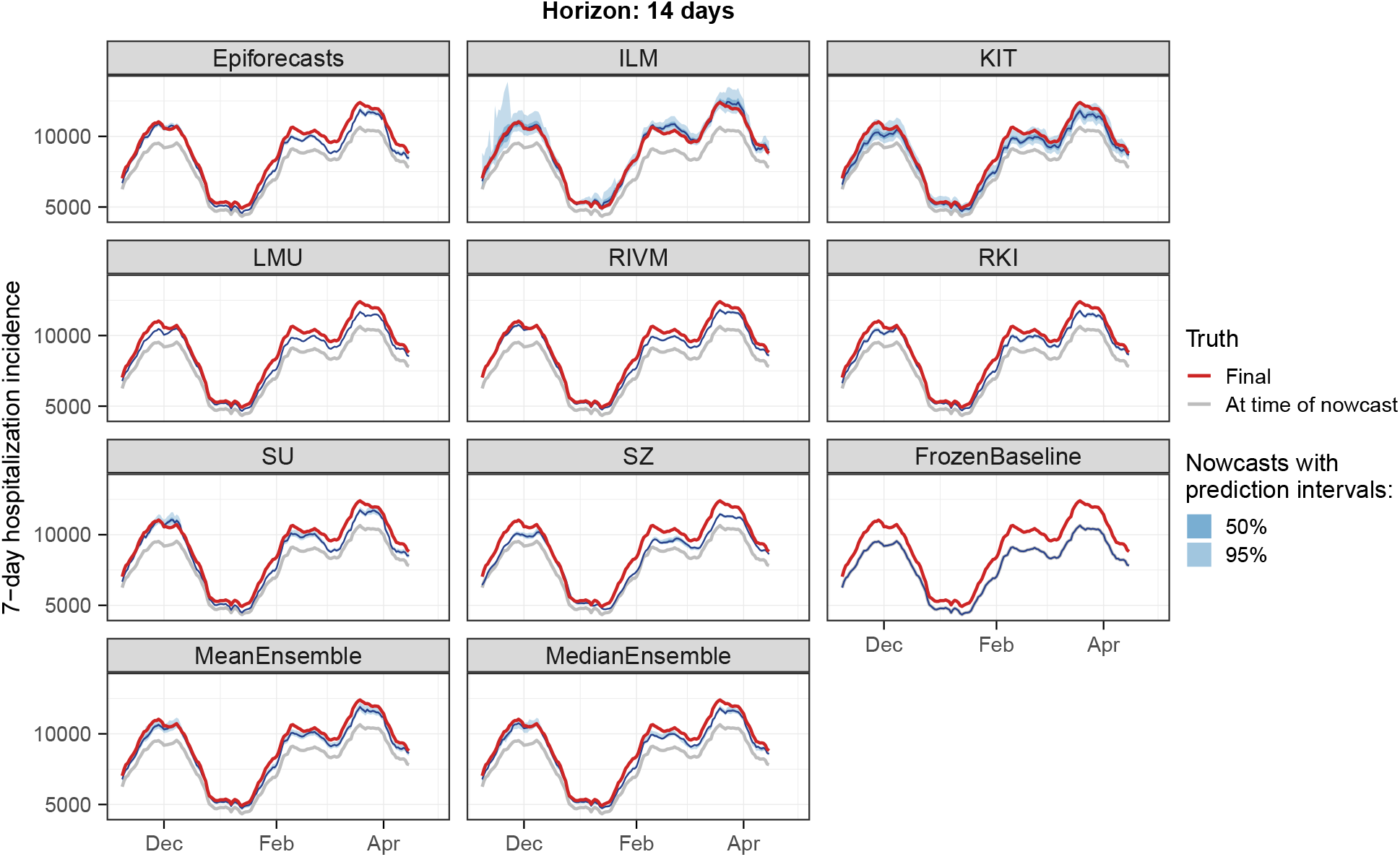
Nowcasts of the 7-day hospitalization incidence as issued 14 days after the respective date (with a horizon of 14 days). Nowcasts are shown for the German national level pooled across all age groups.

### 3.3 Formal evaluation

To consolidate the qualitative findings from the previous section, we turn to a formal evaluation and consider evaluation scores and interval coverage rates. Figure 6 displays the mean and relative WIS values achieved by different models for the three considered aggregation levels (national level, states, and age groups). The left column shows mean scores (on the absolute and relative scale) across all strata and horizons, decomposed into contributions of spread, underprediction, and overprediction. The middle and right panels show the mean WIS and relative WIS by horizon, respectively (−28 to 0 days; see Section 2.5). At the national level and across age groups, the overall scores of the `ILM` model were considerably lower than the scores of all other models. The stratification by horizon indicates that it performed especially well for nowcasts seven or more days back. For the most recent days (−3 to 0 at the national level, −6 to 0 for age groups) the `MeanEnsemble` performed best. Across states, the `MeanEnsemble` outperformed the other models for horizons of −11 to 0 days. For horizons of −28 to −12 days, the `KIT` model achieved the best scores, which (by a narrow margin) led to the best overall result pooled across horizons. The relative scores indicate that pooled over all horizons, most models were able to reduce the error of the uncorrected time series (`FrozenBaseline`) by roughly 80% (relative WIS of 0.2), while the `ILM` model achieved a reduction of about 90% (relative WIS 0.1). It is notable that `ILM` achieved almost constant improvements across horizons, while the improvements achieved by the other models were quite modest for horizons further into the past. To allow for a more detailed exploration of results we provide a display of the distribution of model ranks across individual nowcasting tasks (Supplementary Figure 18) and of scores over time (Supplementary Figure 19). Similarly to [17], we find that the `MeanEnsemble` reliably achieved above-average performance across all locations and age groups (almost never ranking in the bottom).

**Figure 6:**
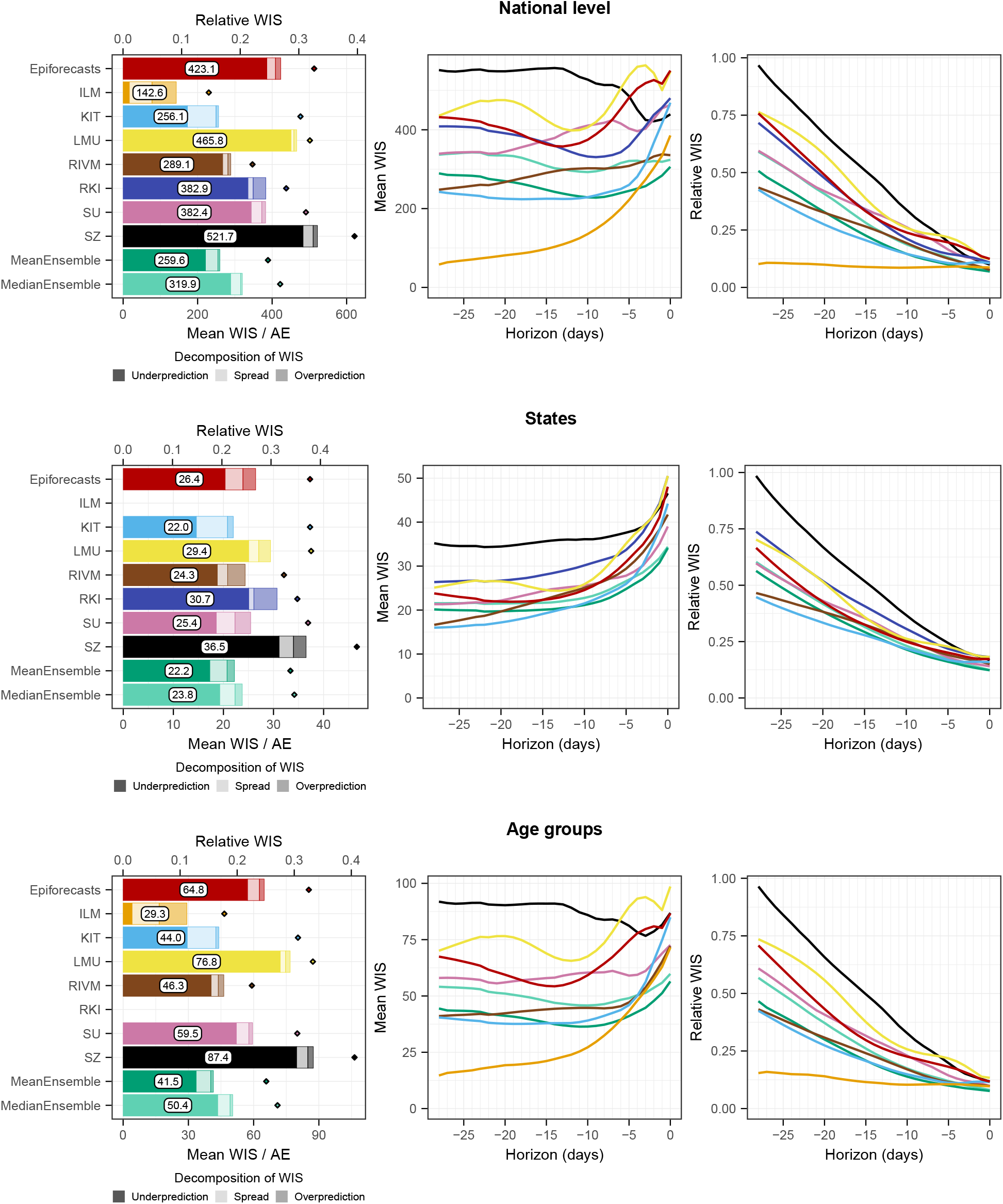
Scores for the national level (top) and averaged across states (middle) and age groups (bottom). The first panel in each row displays the average across all horizons (on the absolute and relative scales). The decomposition into nowcast spread, underprediction, and overprediction (see Section 2.5) is represented by blocks of different color intensities. The absolute error is indicated by a diamond (⋄). The second and third panels in each row show the mean WIS and the relative WIS, respectively, stratified by horizon.

Supplementary Figures 15 and 16 summarize results in terms of mean absolute errors of predictive medians and mean squared errors of predictive means in the same format as in Figure 6. The `ILM` model again performed favorably. Among the remaining models, `RIVM` shows good performance, in many cases outperforming the ensembles. The `KIT` model, on the other hand, which performed relatively well on WIS, achieved below-average results.

Empirical coverage rates of the 50% and 95% prediction intervals are displayed in Figure 7. Results are stratified by aggregation level (national, states, age groups) and nowcast horizon (−28 days to 0 days). The best calibration was achieved by the `ILM` model, with coverage rates close to the nominal levels at most horizons. Only for short horizons of −10 to 0 days coverage dropped moderately. In contrast, the `KIT` model achieved higher coverage rates for horizons between −4 and 0 days, which considerably dropped for nowcasts further into the past. All other models were overconfident and did not reach the respective nominal coverage levels. As for `KIT`, coverage was lower for nowcasts further back in time, for some models to a point where only a few observations were covered at −28 days.

**Figure 7:**
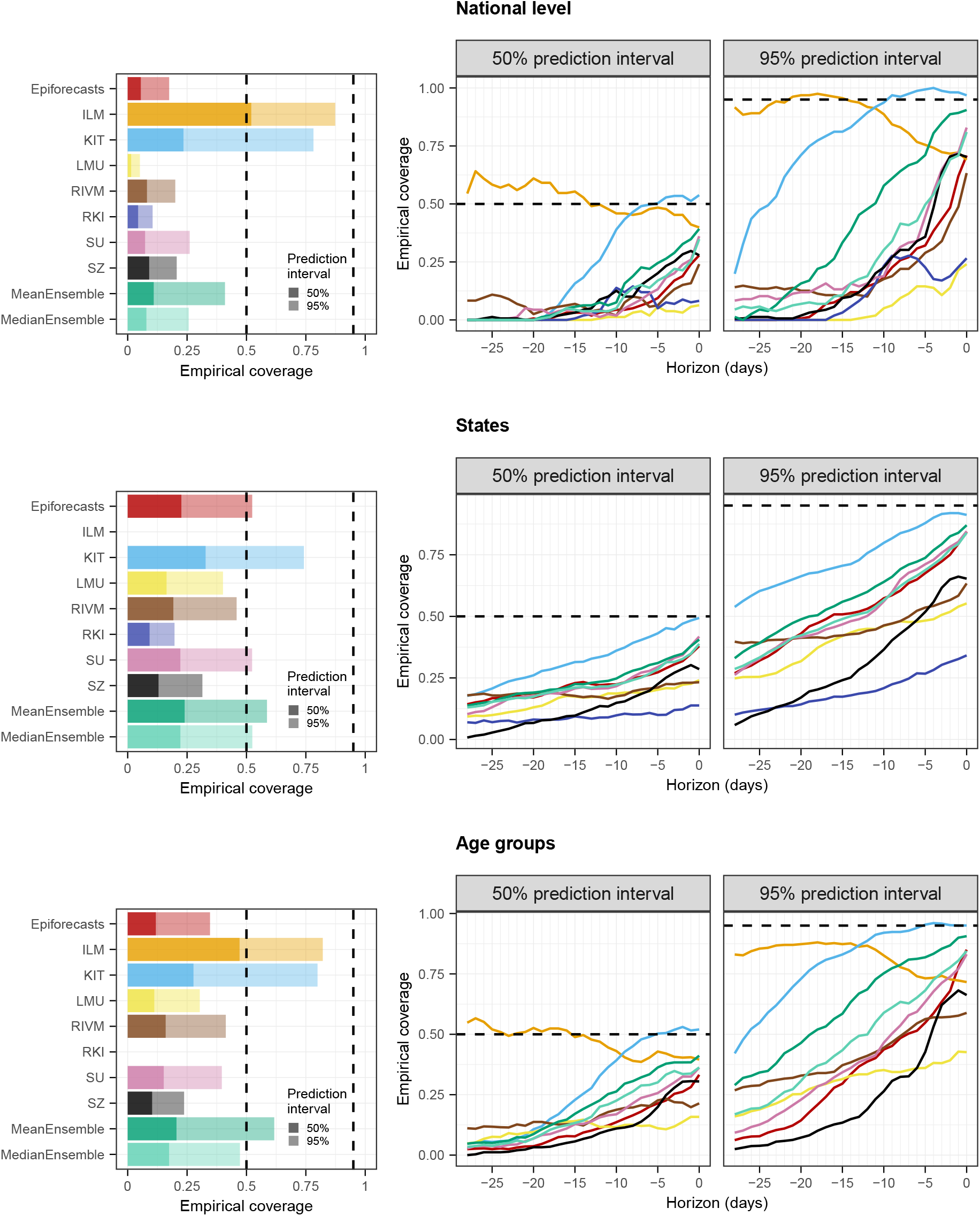
Empirical coverage proportions for the national level (top), across states (middle), and age groups (bottom). The first panel in each row displays the overall coverage of the 50% and 95% prediction intervals across all horizons. The second and third panels in each row show the empirical coverage of the 50% and 95% prediction intervals, respectively, stratified by horizon. The dashed lines indicate the desired nominal levels.

To assess the impact of retrospective fill-in nowcasts for missing submissions, we recomputed relative WIS values using only real-time submissions and the pairwise comparison scheme from [17]. As can be seen from Supplementary Table 6, the results barely change, indicating that fill-in nowcasts did not have a relevant impact on overall scores. Furthermore, as designated in the study protocol, we reran the evaluation using hospitalization incidences per 100,000 population rather than absolute hospitalization counts. The results are displayed in Supplementary Figure 20 and do not differ qualitatively from those in Figure 6.

**Table 6:** Comparison of relative WIS values obtained using retrospective fill-in nowcasts and the pairwise comparison approach from [17] (PC).

|  | National level |  | States |  | Age groups |  |
| --- | --- | --- | --- | --- | --- | --- |
|  | PC | fill-in | PC | fill-in | PC | fill-in |
| Epiforecasts | 0.2679 | 0.2690 | 0.2686 | 0.2686 | 0.2465 | 0.2472 |
| FrozenBaseline | 1.0000 | 1.0000 | 1.0000 | 1.0000 | 1.0000 | 1.0000 |
| ILM | 0.0907 | 0.0907 |  |  | 0.1124 | 0.1117 |
| KIT | 0.1627 | 0.1628 | 0.2227 | 0.2232 | 0.1676 | 0.1677 |
| LMU | 0.2961 | 0.2962 | 0.2993 | 0.2988 | 0.2925 | 0.2926 |
| MeanEnsemble | 0.1643 | 0.1651 | 0.2254 | 0.2254 | 0.1574 | 0.1581 |
| MedianEnsemble | 0.2034 | 0.2034 | 0.2414 | 0.2415 | 0.1919 | 0.1920 |
| RIVM | 0.1845 | 0.1838 | 0.2471 | 0.2473 | 0.1767 | 0.1764 |
| RKI |  | 0.2435 |  | 0.3120 |  |  |
| SU | 0.2427 | 0.2432 | 0.2590 | 0.2585 | 0.2265 | 0.2268 |
| SZ | 0.3314 | 0.3317 | 0.3703 | 0.3709 | 0.3330 | 0.3332 |

<sup>1</sup> No nowcasts submitted for this target.
<sup>2</sup> WIS could only be evaluated for fill-in nowcasts as real-time submissions did not contain all required quantiles.

As we consider the nowcasts for the most recent days the most relevant from a public health perspective, we conclude with an additional non-preregistered summary of scores across horizons −7 to 0 days. Figure 8 shows the average weighted interval scores and interval coverage rates. For this subset of nowcasting tasks, the `MeanEnsemble` outperformed the individual models in all three categories, closely followed by `ILM`. The `KIT` model reaches close to nominal coverage, while the other models are again overconfident.

**Figure 8:**
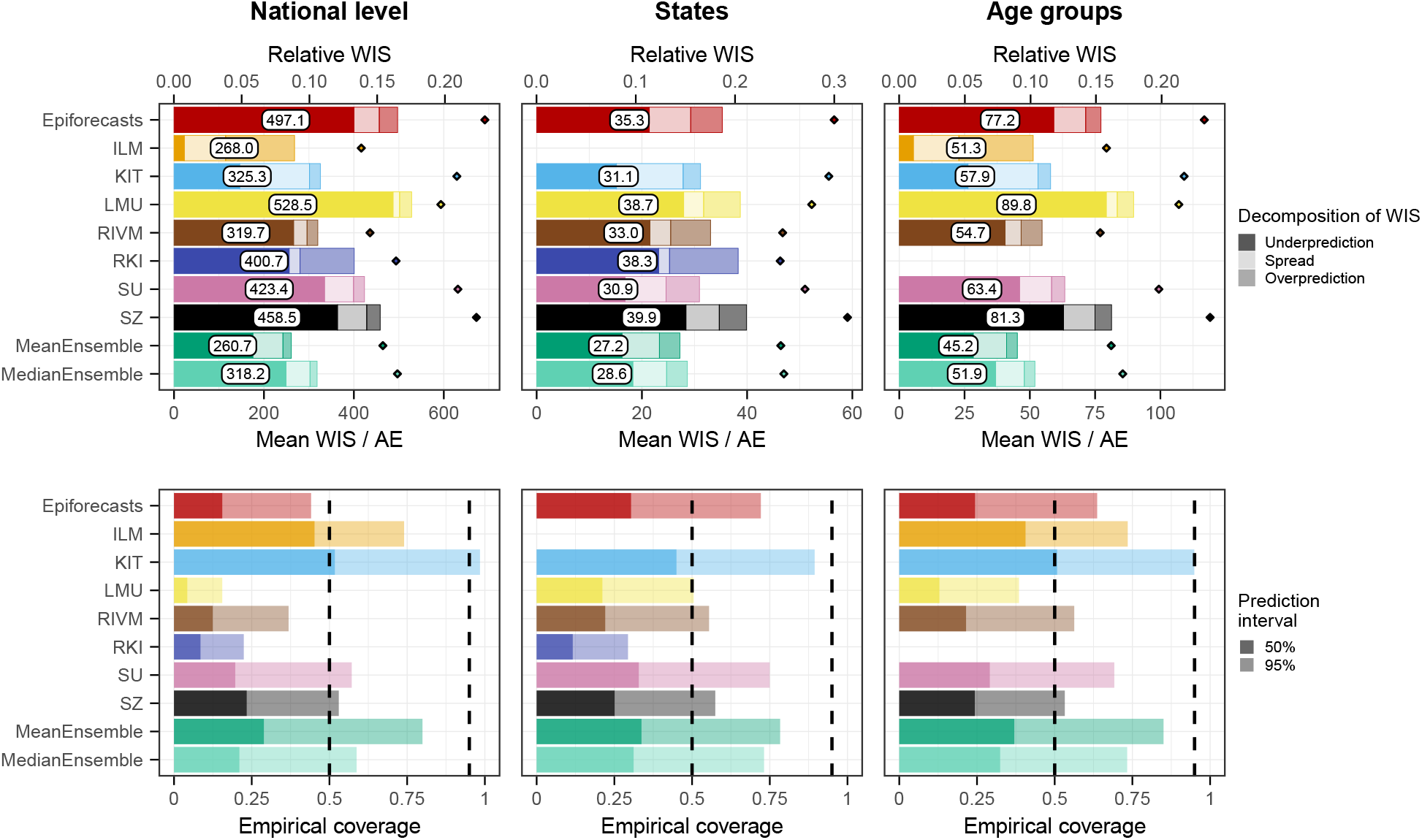
Mean WIS and AE (top) as well as empirical coverage rates (bottom) across horizons from 0 to −7 days.

### 3.4 Interpretation of evaluation results

As some of the presented results may seem contradictory at first sight, we provide some additional interpretations. Firstly, the opposed trends in absolute and relative WIS across horizons in Figure 6 can be interpreted as follows. All nowcasts – including the `FrozenBaseline` – get closer to the later observed final value as time passes and more complete data accumulates; thus, the absolute WIS decreases. However, most models seemed to have more difficulties predicting the small number of late additions than the bulk of early additions, leading to higher relative WIS. A possible explanation is that modelers needed to make a choice on which maximum delay to take into account. In light of Figure 2, the values of around 40 days as chosen by most teams may have been too low and led models to ignore a non-negligible fraction of hospitalizations still to be added. As can be seen from Figure 4, the resulting bias got largely absorbed in the overall uncertainty for same-day nowcasts. For the horizon of −14 days (Figure 5), on the other hand, it caused a visible shift between nowcasts and final values, which likewise led to insufficient coverage of prediction intervals.

The maximum delay chosen may also explain why the `ILM` model, which used a value of 80 rather than 40 days, was the best-performing individual method. However, the model also differed from the others in its general approach, using a regression on case incidences in addition to preliminary hospitalization numbers. We will attempt to shed more light on this aspect in Section 3.6. As a last relevant difference to most other models, the `ILM` approach based uncertainty intervals directly on the errors of past real-time nowcasts, an approach close to the idea of conformal prediction [36]. A similar approach was also taken by the `KIT` model (Appendix E). The fact that these two models achieved the best calibration indicates that this approach may quantify nowcast uncertainty more realistically than standard model-based uncertainty intervals.

The decomposition of the WIS into components for spread, overprediction, and underprediction [21], displayed in the left column of Figure 6, is informative on the challenges the different approaches faced. Penalties for underprediction make up a very large part of the overall scores for all models except for `ILM`. This confirms the observation of a downward bias from Figure 5. The best-performing individual models `ILM` and `KIT` issued predictive distributions with higher variability than the other models, indicated by the larger spread component. As can be seen from the diamond symbols in Figure 15 and in more detail from Supplementary Figure 15, at least the `KIT` model did not issue particularly good predictive medians; instead, its lower WIS values stem from somewhat better uncertainty quantification.

### 3.5 Impact of unusual reporting patterns and changes in virus properties

The nowcasting models in our study assumed either that the probability of hospitalization given a positive test remains roughly constant (the `ILM` model) or that the delay distribution in hospitalizations does so (all other models). In Figure 9, we therefore show four exemplary instances where these assumptions were violated. In mid-November 2022, hospitals in Saxony were overwhelmed [37], leading to disruptions in the reporting system. As a consequence, initial reporting completeness dropped rather suddenly, see also Supplementary Figure 13. This resulted in considerably too low nowcasts, as illustrated in Figure 9a. We note that in this instance we were aware that nowcasts for Saxony were likely unreliable and issued a warning on our website. Figure 9b shows an unusual reporting pattern from the state of Bremen from early 2022. Here, a relevant number of reported hospitalizations got removed from the record on 12 and 13 January, presumably due to faulty initial reporting. Nowcasts issued up to 11 January were thus considerably above the final data version from 8 August. Figures 9c and 9d show issues arising after the Easter weekend of 2022, when initial reporting was considerably lower than usual. As can be seen in Figure 9c, this led to too low ensemble nowcasts on Tuesday, 19 April. Also, over the following days, it seems to have caused issues in the fitting of certain models. As an example, Figure 9d shows the `Epiforecasts` output for Lower Saxony on 20 April. It features an excessively wide prediction interval, likely as a reaction to rapidly shifting delay distributions in the previous days. The `MeanEnsemble`, shown in green, was strongly affected by this unusual behavior of a member nowcast, while the more robust `MedianEnsemble` remained unaffected.

**Figure 9:**
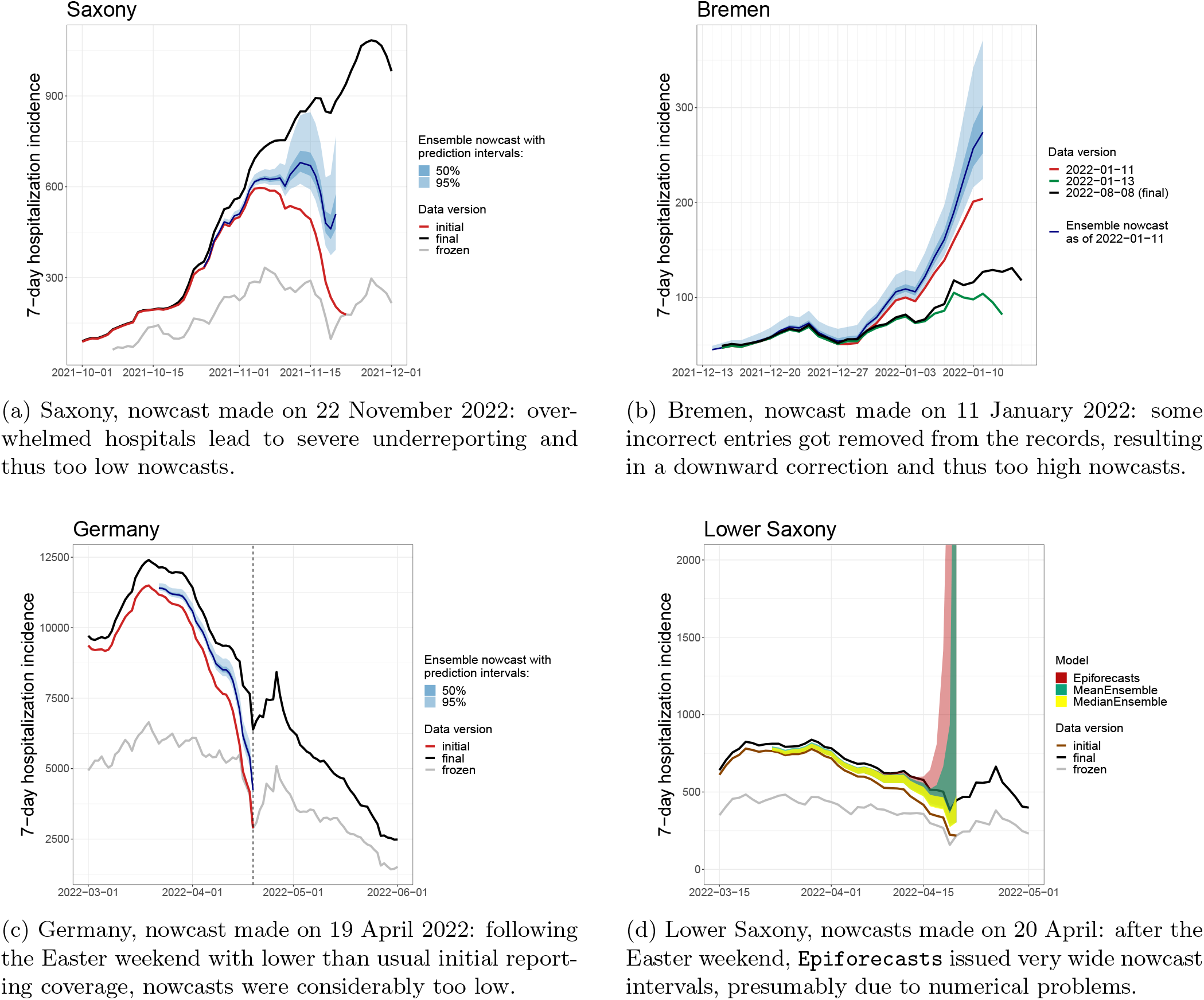
Examples of time points when delay distributions were subject to sudden changes.

A last noteworthy particularity is the behavior of the `ILM` model in January 2022, following the transition from the Delta to the Omicron variant. The Omicron variant is known to have lower clinical severity than the Delta variant [38], meaning that during the transition the ratio of hospitalizations and confirmed cases gradually declined. For the `ILM` model, which assumes this ratio to remain constant, this led to an upward bias in nowcasts, which can be discerned from Figure 4 as an upward bump not present in the other models.

### 3.6 Retrospective variations of models

We next aim to shed some more light on how different modeling choices impact performance and how learnings from our study period facilitated the improvement of methods. To this end, we assess the performance of four variations of previously discussed models, which were applied retrospectively:

- The `LMU` team implemented a new approach to generate uncertainty intervals, which like the `ILM` and `KIT` methods is based on past nowcast errors.
- The `RKI` team obtained nowcasts by aggregating over finer strata and originally assumed independence across strata to generate prediction intervals at the aggregate level. This was changed to an assumption of strong correlations across strata, leading to wider prediction intervals.
- The `KIT` team reran its model with an increased maximum delay of 80 days, in contrast to the 40 days used in real time. This also required an increased length of training data, which was set to 100 days.
- Conversely, the `ILM` model was rerun with a maximum delay of 42 rather than 84 days, which is comparable to the maximum delays used by the remaining models. This was not meant as an improvement but as an adjustment to assess the impact of longer/shorter maximum delays.

The results are shown in Figure 10. They indicate improvements across all aggregation levels and horizons for the revised `LMU`, `RKI`, and `KIT` models. In particular across age groups, the `KIT` model now came close to the performance the `ILM` model achieved in real time. The coverage proportion of prediction intervals was increased for all three models, with the updated `KIT` model even leaning towards over-coverage (too wide intervals). The `LMU` and `RKI` models, on the other hand, remained overconfident. Decreasing the maximum delay in the `ILM` model slightly reduced the overall performance on the national level, while average scores across age groups remained almost unchanged. The score decomposition shows that the adjusted model tended to underpredict (similarly to the other models), while the original model tended to overpredict. Possible explanations will be discussed in Section 4.

**Figure 10:**
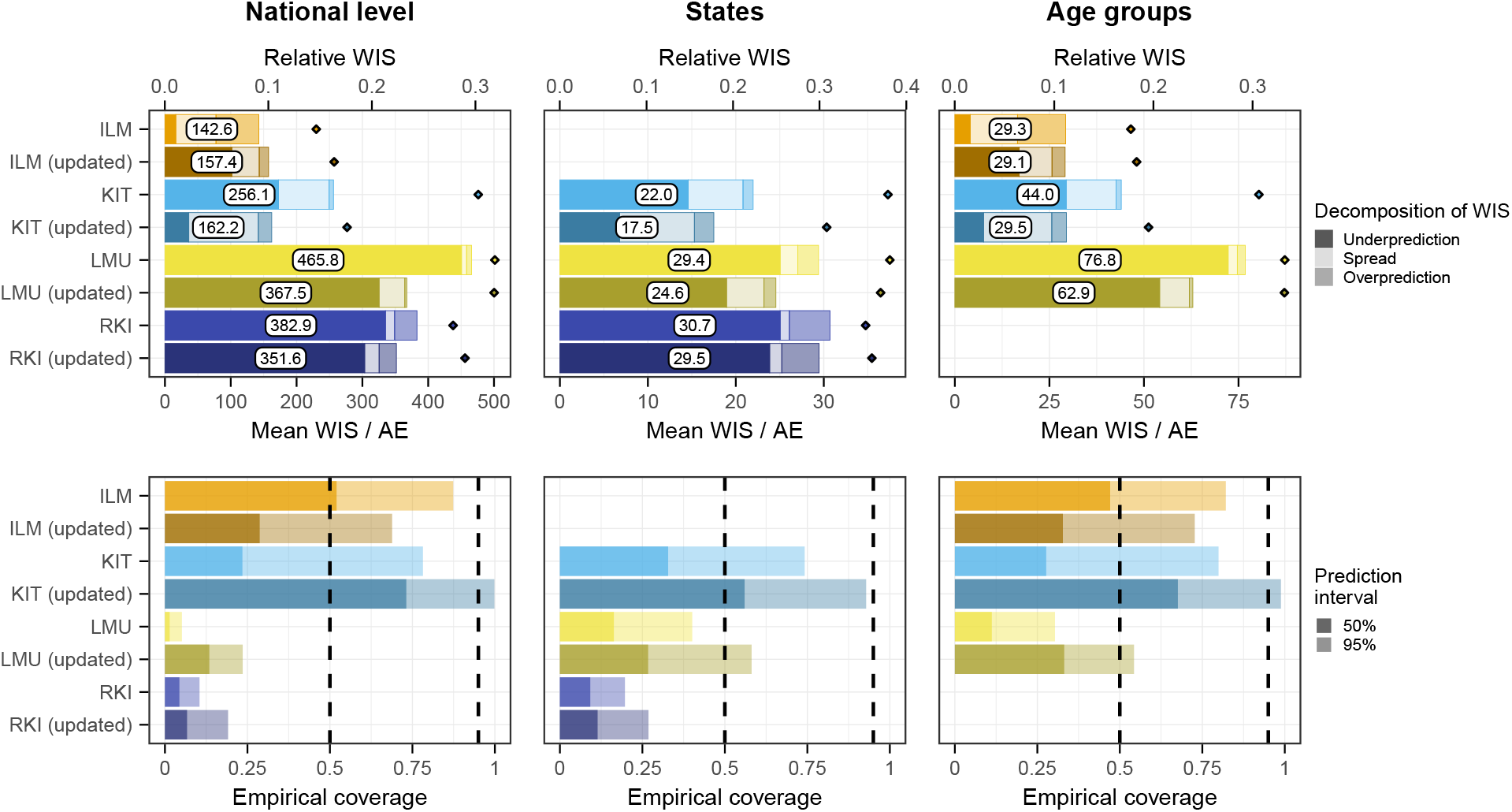
Comparison of retrospective variations of the `ILM`, `KIT`, `LMU`, and `RKI` models and the same models as submitted in real time. Results are comparable to those from Figures 6 and 7.

### 3.7 Sensitivity of results to definition of final data

In our study protocol, we specified that the final state of the time series to be predicted was the version available on 8 August 2022, i.e., 100 days after the end of the study period. However, as we became aware of the fact that data revisions could occur with considerably longer delays than initially expected, we performed a sensitivity analysis to assess the impact of this choice. Figure 11 shows how the average WIS aggregated over horizons and different levels of stratification (i.e., the results shown in the left column of Figure 6) change when using a different data version as the final one. It can be seen that the average scores of all models except for `ILM` increase in parallel the later a data version is used, with a slightly flatter increase for `KIT`. This is because these models tend to underpredict, and as time passes and more additions are made to the data, this problem is exacerbated. For the `ILM` model, which tends to overpredict, average scores initially decline and then plateau, leading to an even more pronounced lead relative to the other models. As can be seen from Supplementary Figure 17, using a later data version for evaluation, `ILM` ultimately also surpasses the ensemble when restricting results to horizons 0 to 7 days back.

**Figure 11:**
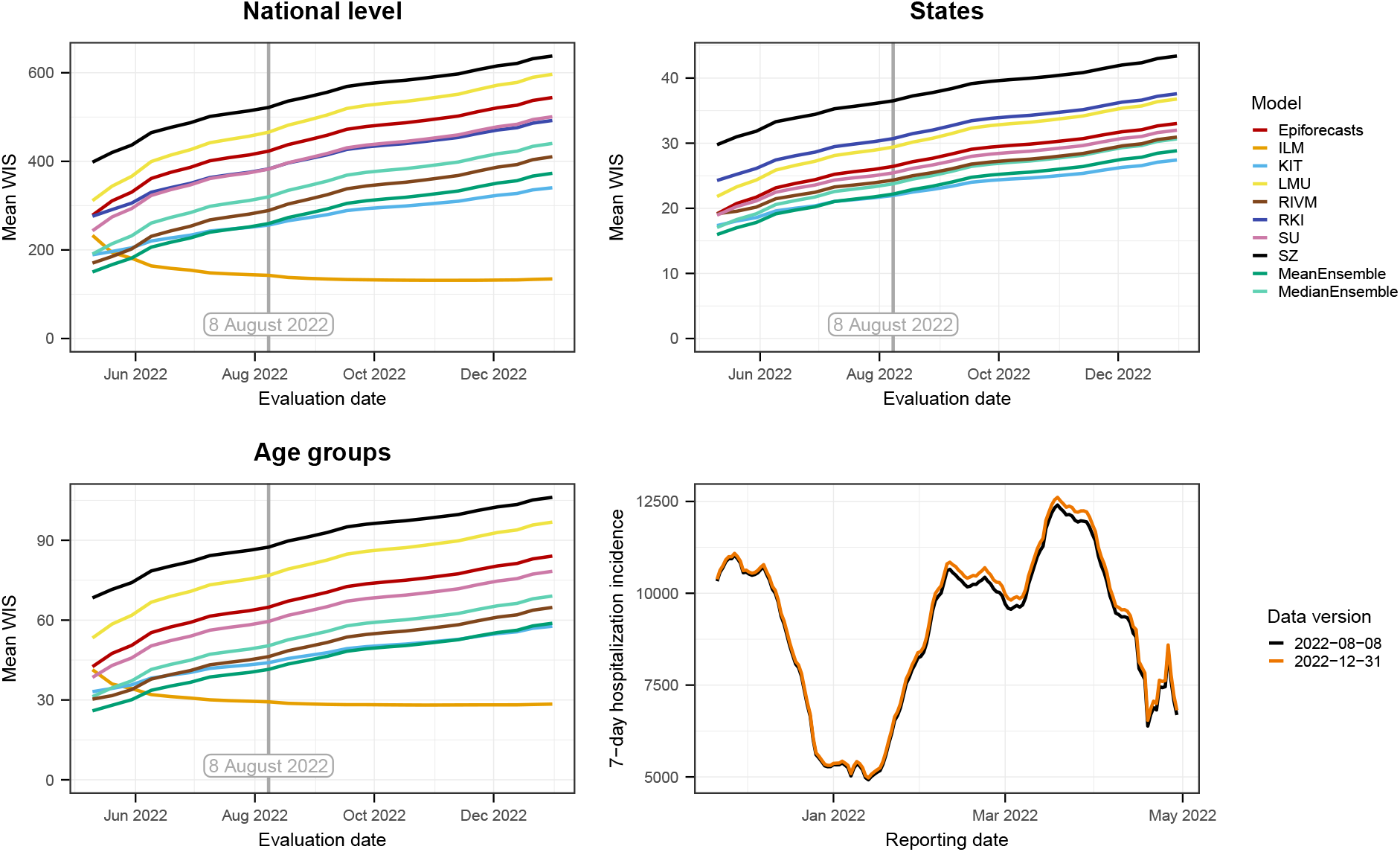
Mean WIS of the compared models computed using different data versions as the “final” version. The version prespecified in the study protocol is 8 August 2022, marked by a vertical line. Top left: national level. Top right: averaged over states. Bottom left: averaged over age groups. The bottom right panel overlays the national-level data versions of 8 August and 31 December to illustrate the importance of late revisions.

An alternative to choosing one specific data version as the nowcast target is a “rolling” approach that considers delayed reports for each reference date up to a specified maximum delay. Figure 12 shows the results this approach yields for a maximum delay of 40 days, which corresponds roughly to the maximum delay used by most teams. As this target definition is better aligned with the practical implementations teams chose, it is not a surprise that mean WIS values are lower and coverage is higher. The `ILM` model (in its adjusted version with a maximum delay of 42 days) now shows quite similar performance to the other approaches, with a tendency to overpredict. The score components for the other models are more balanced and the ensemble nowcasts clearly lead the field. Retrospectively, we think that this definition of targets might have been a more coherent and operationally meaningful approach, see Section 4 for a discussion.

**Figure 12:**
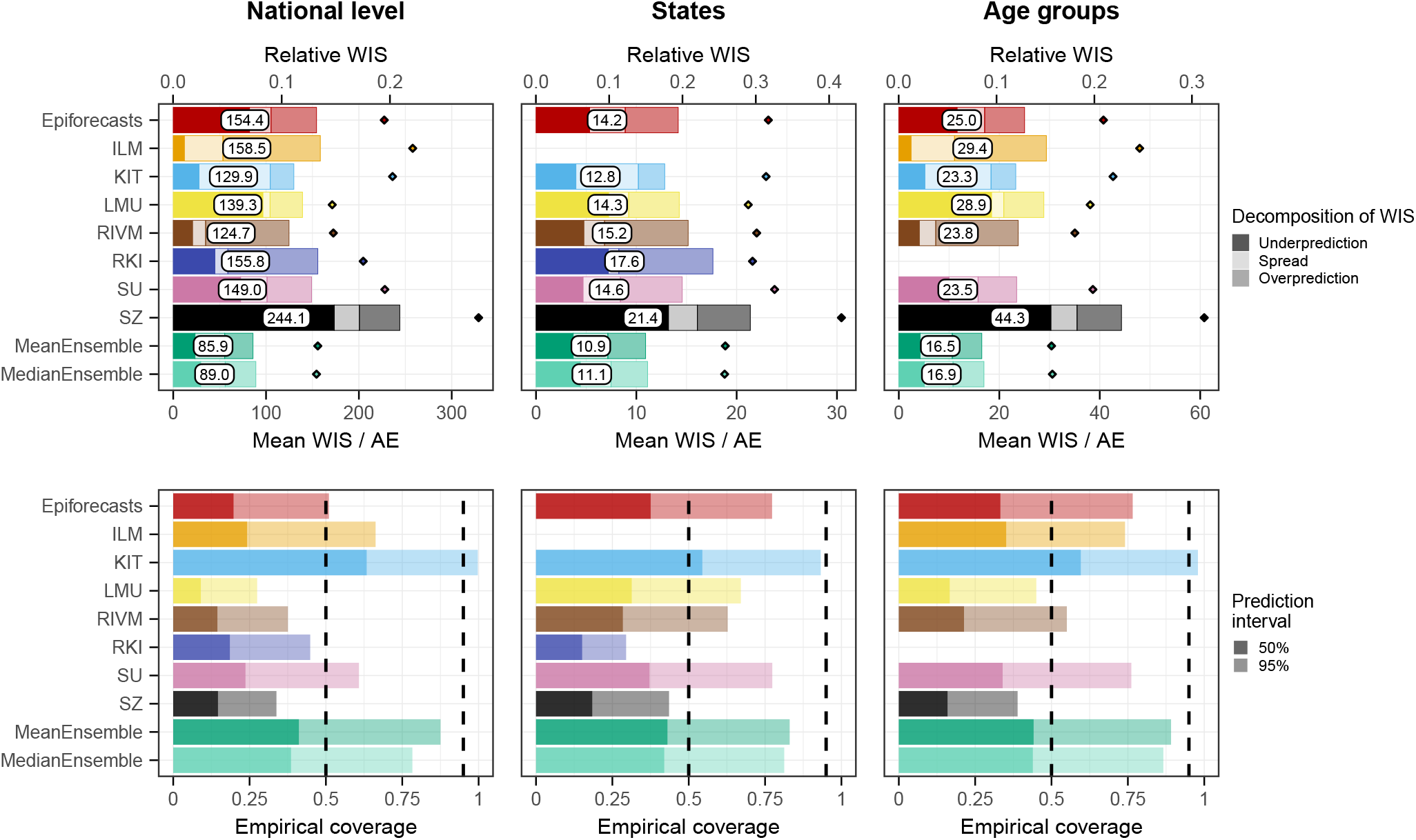
Mean WIS and coverage of nowcasts with respect to a revised target defined as the number of hospitalizations reported with a delay of up to 40 days. For the `ILM` model we used the revised model with a maximum delay of 42 days and also recomputed the ensembles with these revised nowcasts. For the other models, the assumed maximum delays are approximately aligned with the redefined target, see Table 1.

**Figure 13:**
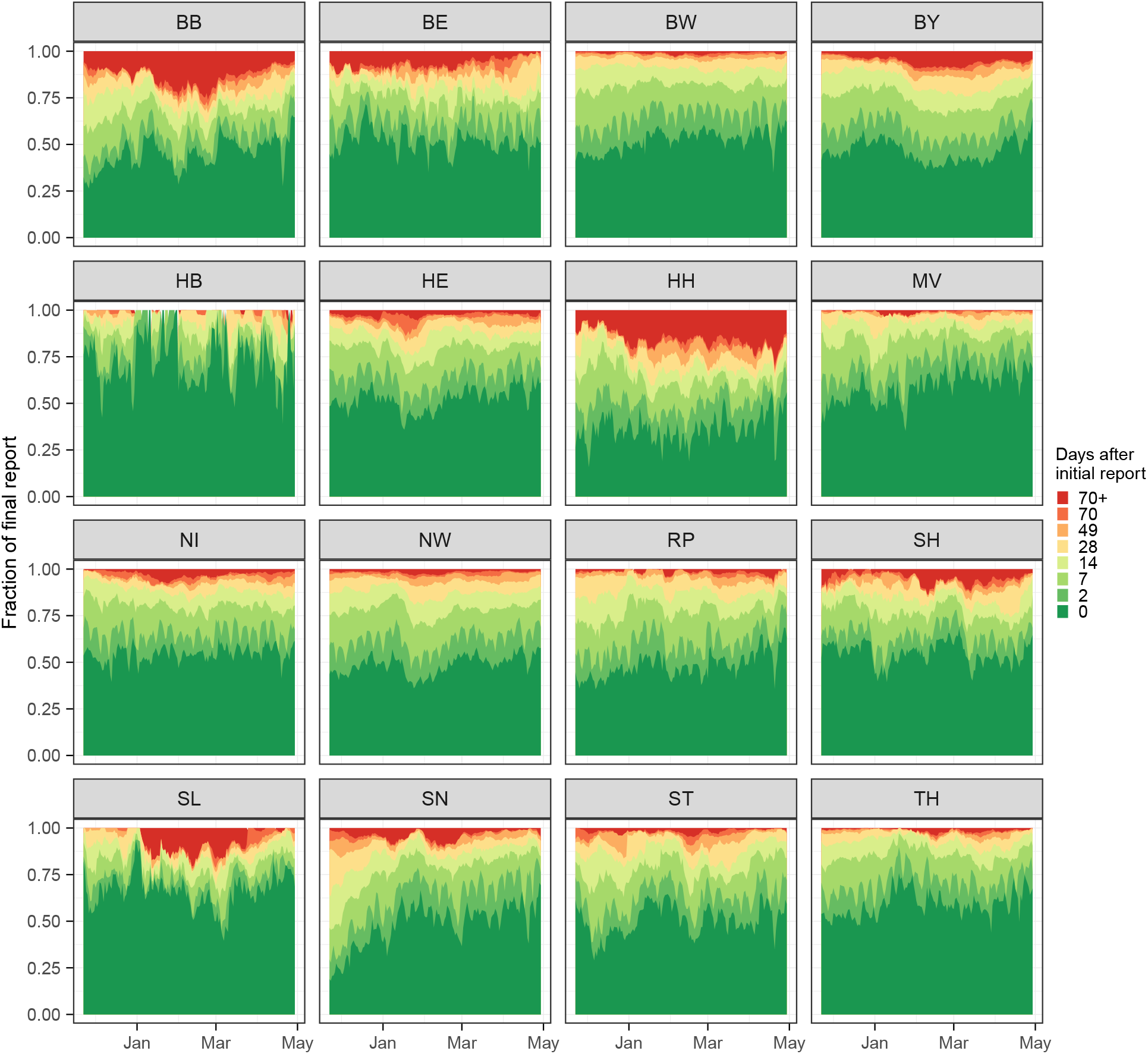
Completeness of 7-day hospitalization incidences 0 to 70 days after the respective reference date. Temporal development in the 16 German states. Abbreviations of federal states: BB = Brandenburg, BE = Berlin, BW = Baden-Württemberg, BY = Bavaria, HB = Bremen, HE = Hessen, HH = Hamburg, MV = Mecklenburg-Vorpommern, NI = Lower Saxony, NW = North Rhine-Westphalie, RP = Rhineland Pallatinate, SH = Schleswig Holstein, SL = Saarland, SN = Saxony, ST = Saxony Anhalt, TH = Thuringia.

**Figure 14:**
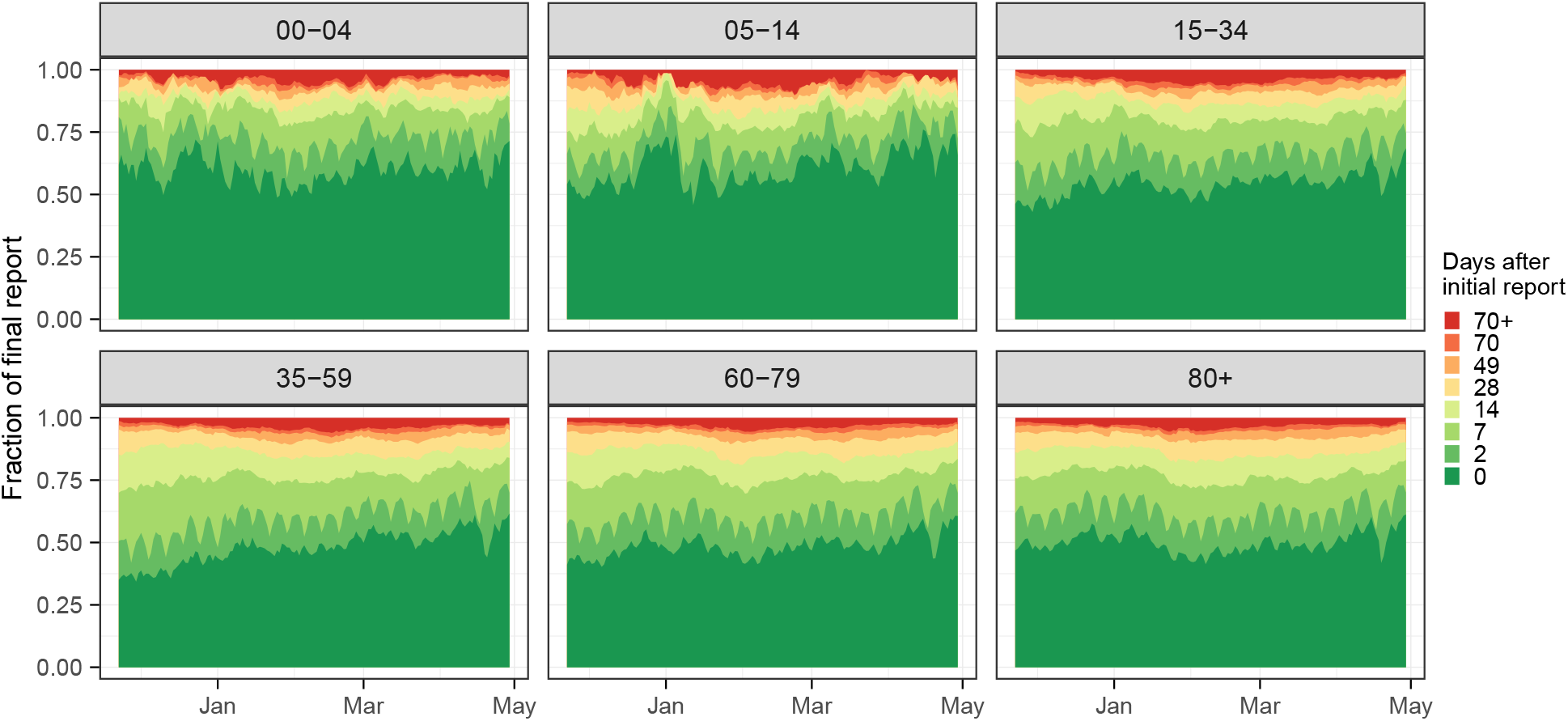
Temporal development of reporting completeness in different age groups.

**Figure 15:**
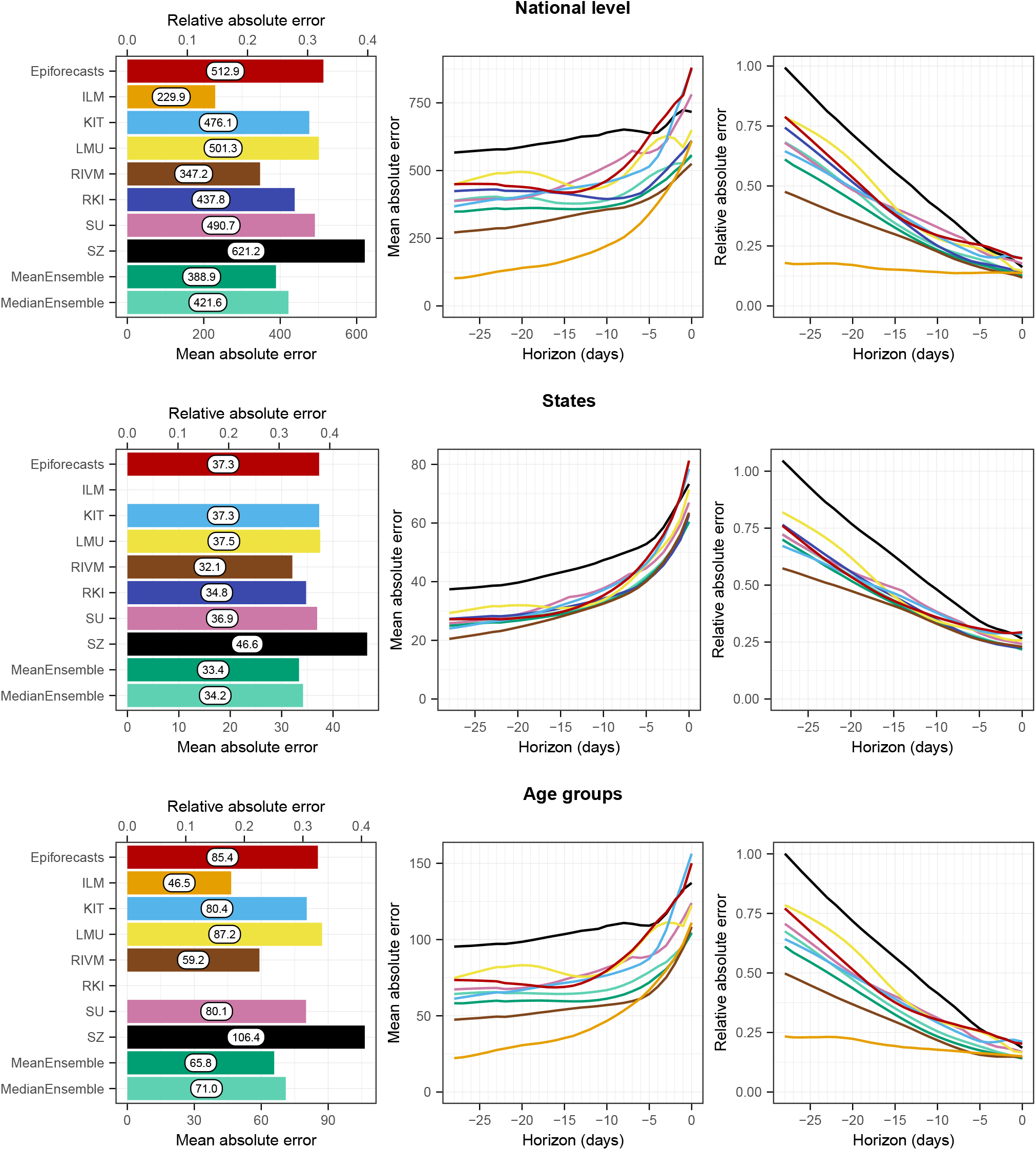
Mean absolute errors for the national level (top) and averaged across states (middle) and age groups (bottom). The first panel in each row displays the average across all horizons (on the absolute and relative scales). The second and third panels in each row show the mean MAE and the relative MAE, respectively, stratified by horizon.

**Figure 16:**
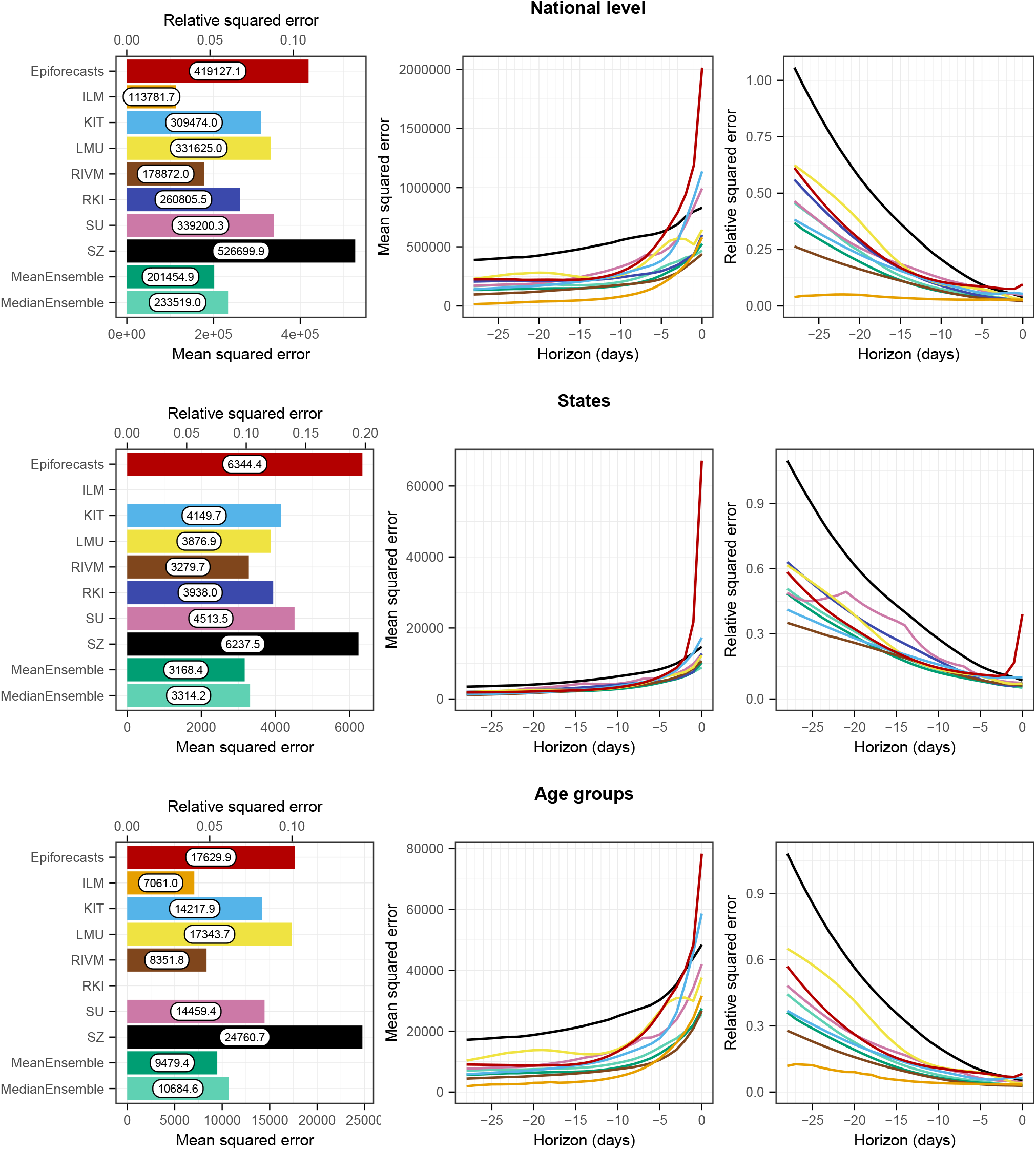
Mean squared errors for the national level (top) and averaged across states (middle) and age groups (bottom). The first panel in each row displays the average across all horizons (on the absolute and relative scales). The second and third panels in each row show the MSE and the relative MSE, respectively, stratified by horizon.

**Figure 17:**
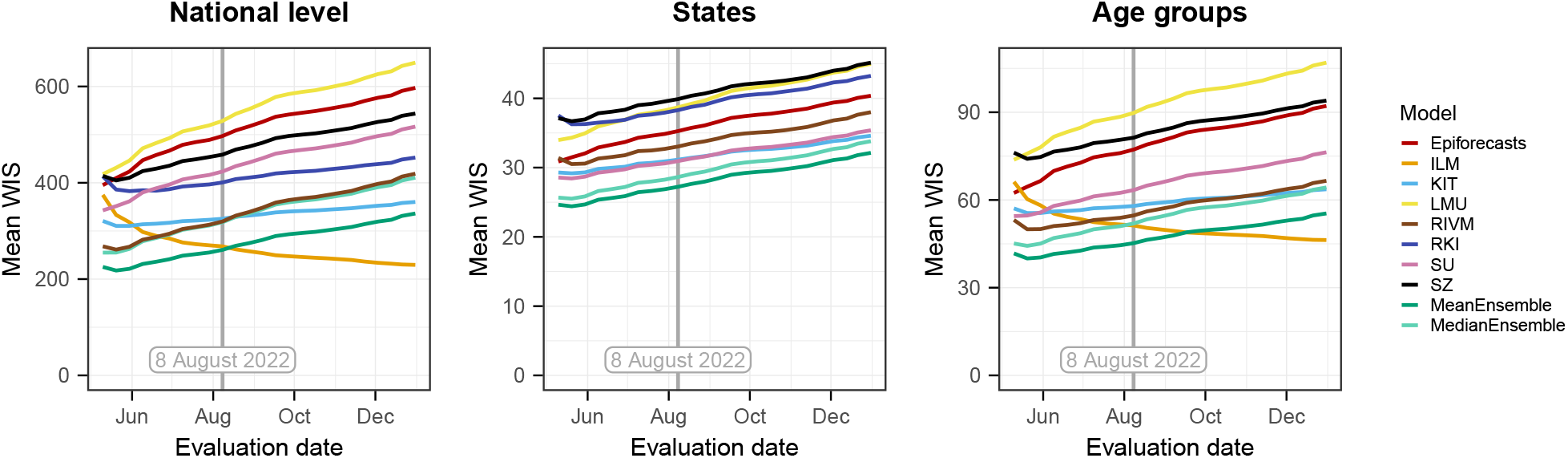
Mean WIS across horizons from 0-7 days of the compared models computed using different data versions as the “final” version. The version prespecified in the study protocol is 8 August 2022, marked by a vertical line.

**Figure 18:**
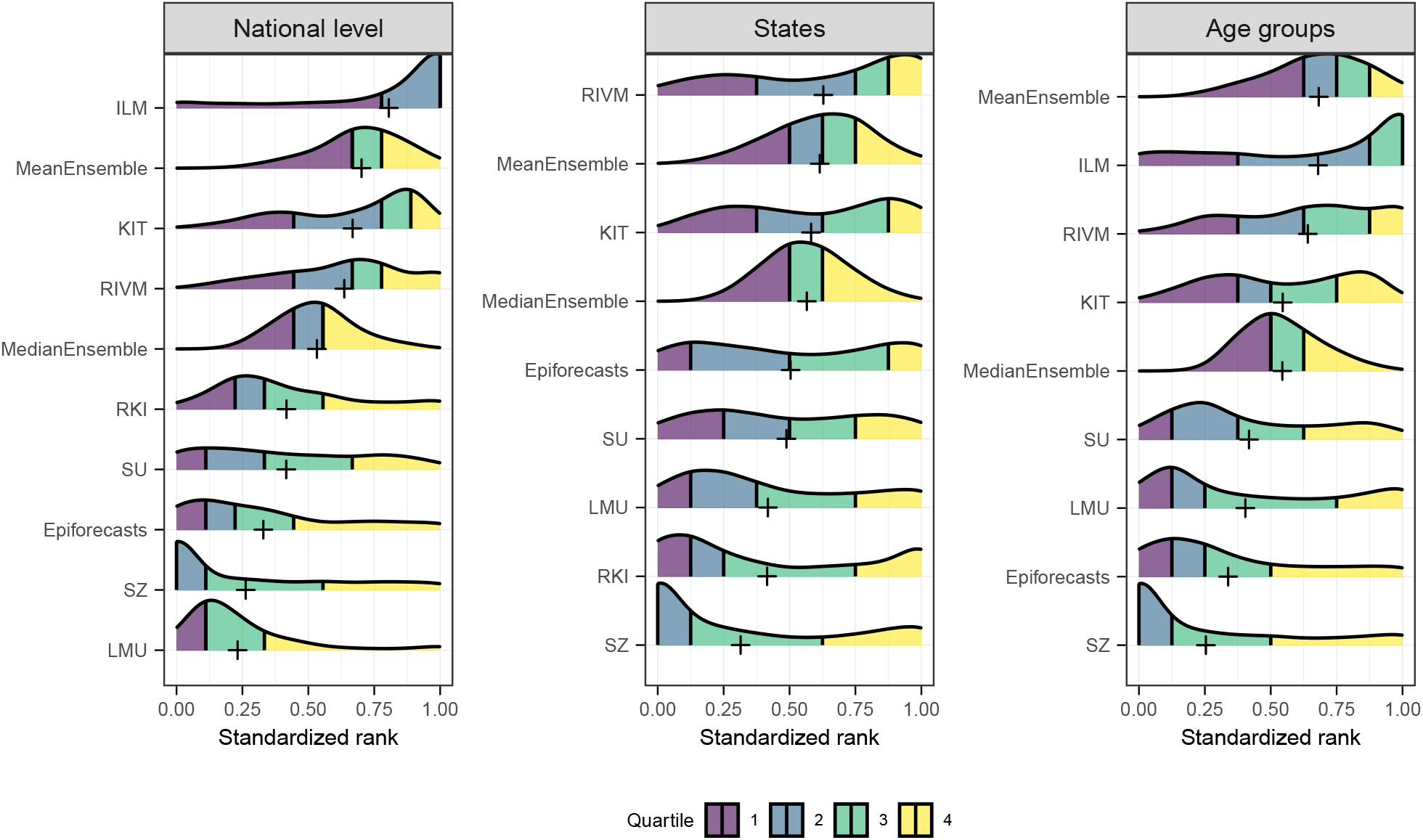
Distribution of each model’s standardized rank for each nowcast-observation pair (see [17] for details on the definition) The models are ordered by the mean standardized rank, which is indicated by a plus sign.

**Figure 19:**
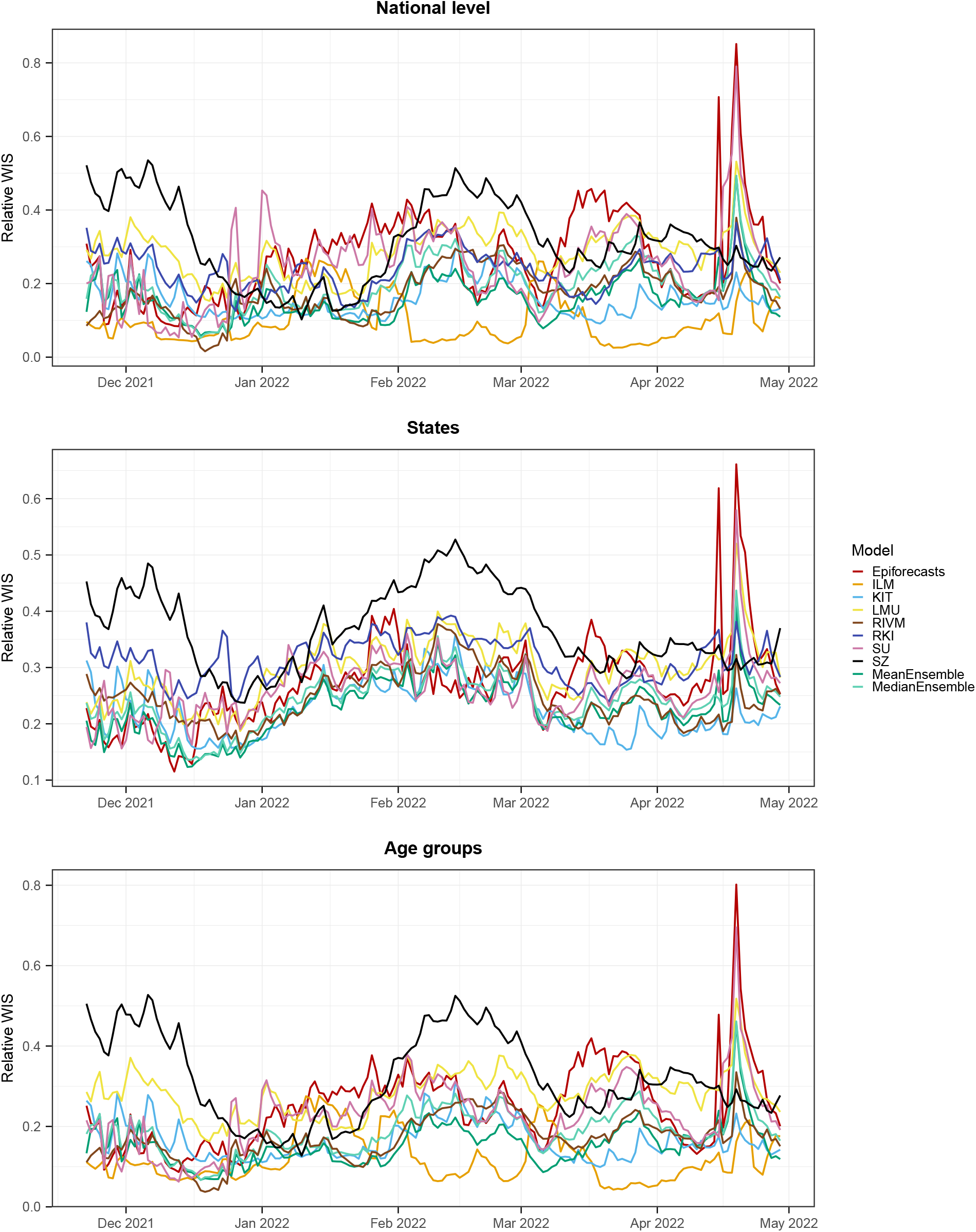
Relative mean WIS across all horizons by nowcast date and stratification level. Top: national level; middle: across states; bottom: across age groups.

**Figure 20:**
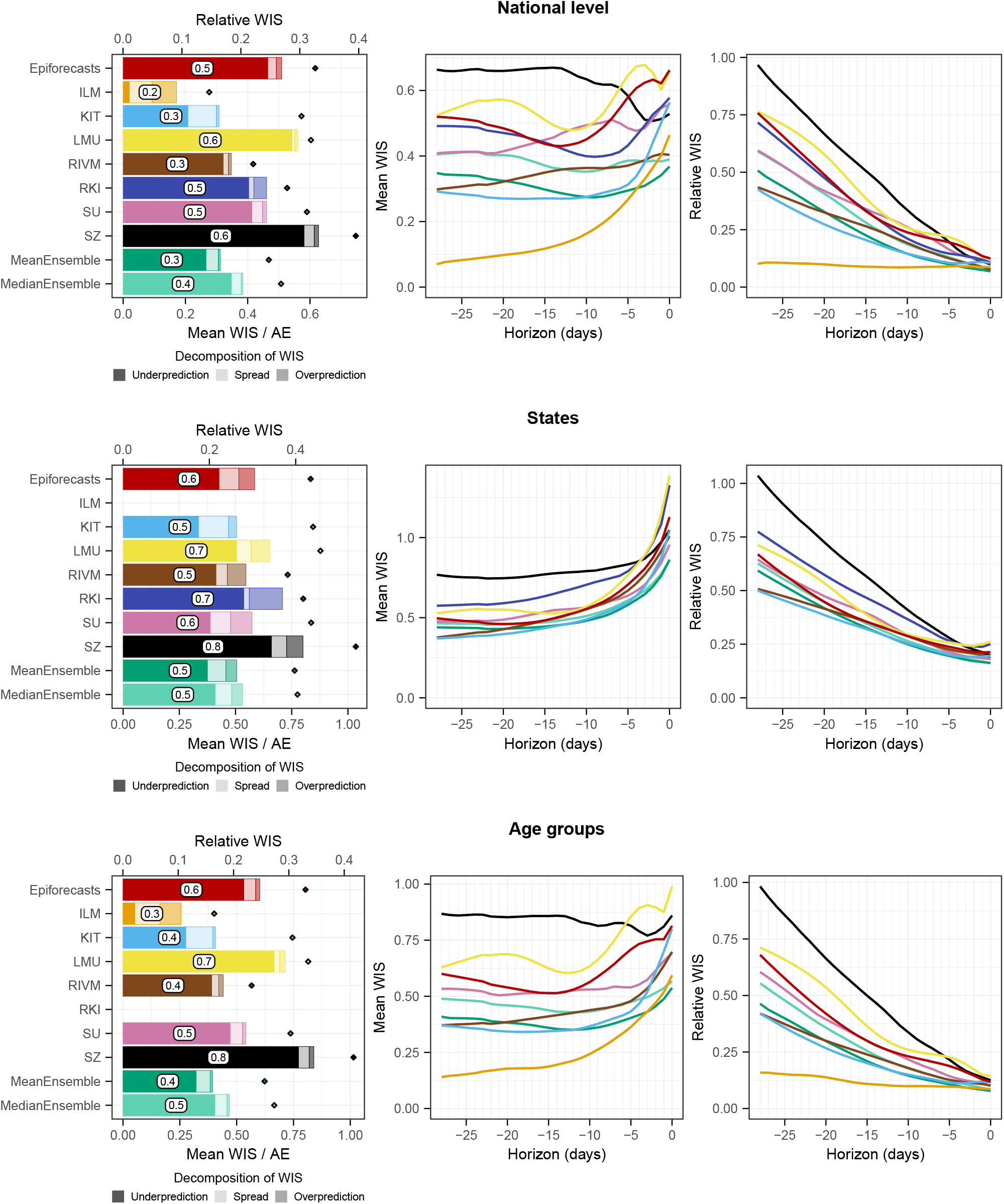
Scores as in Figure 6, but for hospitalizations by 100,000 inhabitants. This gives similar weight to all states irrespective of their population size.

## 4 Discussion

In this paper, we presented results from a preregistered study to evaluate probabilistic real-time nowcasts of the 7-day hospitalization incidence in Germany from November 2021 to April 2022. We found that all models were able to correct for a large part of the biases caused by reporting delays. Further, we identified calibration of uncertainty intervals as a major challenge, as the empirical coverage rates achieved by most models were considerably below the respective nominal levels. The exception was the `ILM` model which also stood out in terms of score-based performance for the national level and across age groups. Reasons for insufficient coverage likely include too inflexible modeling of dispersion and delay distributions, but also the fact that most teams truncated delay distributions at a too short maximum delay.

Our analyses from Sections 3.6 and 3.7 suggest that the success of the `ILM` model arose from the interplay of two aspects. On the one hand, it used a maximum delay that is longer than those of the other models, but, judging by Figure 2, still somewhat too short. On the other hand, the model appears to have a tendency to slightly overpredict the number of hospitalizations added up to a given maximum delay. As these two aspects work in different directions, the resulting nowcasts are overall well-aligned with the defined target (data version from 8 August 2022). Whether nowcasts taking into account case incidence inherently perform better needs to be explored in future work. A combination of different data streams may reduce the dependence of nowcasts on certain assumptions, such as the constant completeness of initial reports. The `Epiforecasts` and `SU` models have been extended in this direction. In an application to COVID-19 deaths in Sweden [39], the inclusion of reported cases and intensive care admissions as leading indicators indeed led to improved predictions.

The `MeanEnsemble`, along with the `KIT` model, performed best across federal states (for which the `ILM` model did not provide nowcasts). Also, it showed very good relative performance for horizons −7 to 0 days, as well as for the revised “rolling” target. We therefore conclude that ensemble approaches are a promising avenue in order to improve disease nowcasting. However, our case study also illustrates a limitation of unweighted ensembles. The ensemble may have been imbalanced in the sense that a majority of its members followed similar strategies and had similar weaknesses (specifically a downward bias due to neglecting very long delays). A weighted ensemble could have capitalized on the strengths of the `ILM` model, which followed a conceptually different approach and could have served as a counterbalance. However, it is not obvious how ensemble members can be assigned weights in real time in a nowcasting setting. This represents an interesting future research area.

A difficulty we encountered in terms of our study design is that results depend on which data version is used as the “final” one (i.e., the values against which nowcasts are evaluated; Section 3.7). As the choice of 8 August 2022 was preregistered and known to all participating teams, the prediction task was well-defined, and we stuck to this choice for our main analysis. Nonetheless, this definition, which was based on the assumption that data would be stable after 100 days, turned out not to be ideal in retrospect. In particular, it implies that for the first day of our study period (22 November 2021), retrospective additions could accumulate over 259 days, while for the last (29 April 2022) this was restricted to 100 days. Defining the nowcast target in a “rolling” fashion as explored in Section 3.7 might have been a more appropriate choice. This would have been a more clearly defined modeling task, and modelers would not have had to choose a maximum delay for their models themselves.

The question of whether additions should be ignored from a certain maximum delay onward is closely linked to what these additions actually mean and whether they are relevant from a public health perspective. As mentioned in Section 2.1, the 7-day hospitalization incidence also contains hospitalizations that are not primarily due to COVID-19. These hospitalizations have been found to represent a considerable fraction [40]. It seems plausible that very long delays are due to large time differences between the positive test and hospital admission, in which case the share of hospitalizations that are not primarily due to COVID-19 may be high. Also, it can be questioned whether hospitalizations a long time after a positive test are relevant for the real-time assessment of healthcare burden. Both aspects strengthen the case for limiting nowcasting to hospitalizations reported up to a carefully chosen maximum delay.

The definition of the *frozen* values used by the Robert Koch Institute when applying legally defined thresholds can be seen as a strong form of discarding delayed hospitalizations. It has the advantage of simplicity and unambiguity, which are required for actionable guidelines in a legal context. After all, it seems difficult to integrate complex statistical methods with many tuning parameters into a binding legal document. An important downside, however, is that the same *frozen* value can mean rather different things at different time points and in different locations, due to differences in initial reporting completeness. We thus argue that outside of purely legal considerations, nowcasts can provide a more thorough picture of current developments.

All nowcasts generated within the presented collaborative project are available in a public repository (see data availability statement). Time-stamped versions of hospitalization data as available at different points in time can be retrieved from the commit history of the repository as well as directly from Robert Koch Institute [23]. We hope that this data can be of use as a benchmarking system for future nowcasting methods. In this context, we note, however, that the present paper is a comparison of *nowcasting systems*, which are given by a statistical model, but also various additional analytical choices, in particular the assumed maximum delay and the length of training data used at each time point. These decisions can have a substantial impact on predictive performance (see Section 3.6) and are easier to get right in hindsight than in real time. To ensure a fair comparison, it may therefore be reasonable to use the “rolling” target as discussed in Section 3.7.

The nowcasts produced for this project were routinely displayed by numerous German-speaking media, including *Die Zeit, Neue Zürcher Zeitung* and *Norddeutscher Rundfunk*. While some displays were limited to the point nowcasts (predictive medians), others made the predictive uncertainty clearly visible. This development should be further encouraged by scientists advising the media on the display of epidemiological data and models. In this context, we also note that data journalists were overall hesitant to use the ensemble nowcasts and prioritized individual nowcasts based on methods described in peer-reviewed publications. Interestingly, the best-performing models in our study were the `MeanEnsemble` and the yet unpublished `ILM` approach. However, our analyses show that all compared approaches provided a good qualitative impression of current incidence trends and levels, and we consider each of them a helpful addition and improvement over showing uncorrected data.

To conclude, we highlight some advantages of the collaborative nowcasting approach adopted in our study. The ensemble nowcast not only showed strong relative performance but was also the most consistently available nowcast, with almost all other models unavailable due to technical problems on some days during the study period. Additionally, our collaborative approach fostered frequent exchange and interaction among modelers via bi-weekly coordination calls, creating a valuable platform for knowledge sharing, feedback, and collaboration on methodological advancements. Through these interactions, the project facilitated model improvements, as seen for the `LMU`, `RKI` and `KIT` approaches in Section 3.6, and fostered discussion on new methodological topics beyond the scope of the present article. For example, the *Epinowcast community* (https://www.epinowcast.org/) was established to build and assess real-time analysis tools, publically available in the R package `epinowcast` [41]. The benefits of our collaborative approach demonstrate the importance of ongoing communication and cooperation in the development and refinement of epidemiological models, particularly during rapidly evolving public health crises such as infectious disease outbreaks.

## Data Availability

The nowcasts collected for this study are available in a GitHub repository (https://github.com/KITmetricslab/hospitalization-nowcast-hub), with a stable release published at https://zenodo.org/record/7828604. The repository also contains the truth data used for evaluation. An interactive dashboard to visualize the nowcasts is provided at https://covid19nowcasthub.de/.
Code to reproduce results and figures are provided at https://github.com/dwolffram/hospitalization-nowcast-hub-evaluation. A list of the participants' code repositories can be found in Appendix D.

https://github.com/dwolffram/hospitalization-nowcast-hub-evaluation

## Data availability

The nowcasts collected for this study are available in a GitHub repository (https://github.com/KITmetricslab/ hospitalization-nowcast-hub), with a stable release published at https://zenodo.org/record/7828604.

The repository also contains the truth data used for evaluation. An interactive dashboard to visualize the nowcasts is provided at https://covid19nowcasthub.de/.

## Code availability

Code to reproduce results and figures are provided at https://github.com/dwolffram/hospitalization-nowcast-hub-evaluation. A list of the participants’ code repositories can be found in Appendix D.

## Acknowledgements

We would like to thank Tilmann Gneiting for his helpful comments and feedback on the evaluation approach. J. Bracher and M. Schienle acknowledge support from the Helmholtz Foundation via the projects *SIM-CARD* and *COCAP*. J. Bracher, D. Wolffram and M. Schienle were moreover supported by the German Federal Ministry of Education and Research (BMBF) via the project RESPINOW. J. Bracher’s work has been partly funded by the Deutsche Forschungsgemeinschaft (DFG, German Research Foundation) – project number 512483310. D. Wolffram is grateful for support from the Klaus Tschira Foundation. D. Wolf-fram’s contribution was moreover supported by the Helmholtz Association under the joint research school HIDSS4Health – Helmholtz Information and Data Science School for Health. S. Abbott and S. Funk were supported by The Wellcome Trust (210758/Z/18/Z). F. Günther was supported by NordForsk (grant no. 105572).

## Appendix

### A Deviations from study protocol and completeness of nowcasts

As we have deviated in some minor parts from the study protocol, we provide a list of these adjustments.

### B Supplementary Figures

### C Sensitivity analysis via pairwise comparisons

#### C.1 Motivation and procedure

In some instances, teams failed to submit nowcasts in time and had to fill them in post hoc. Allowing them to do so may seem lenient as, in principle, teams could use additional information, thus unfairly improving their nowcasts. As specified in our protocol, we thus perform a sensitivity analysis based purely on nowcasts submitted on time of the respective day.

As each team failed to submit nowcasts on different days this leads to a setting where methods need to be compared based on incongruent sets of nowcasting tasks. In this setting, the relative WIS could still be evaluated for each method by considering only the subset of targets treated by the respective method.

This, however, ignores that improving upon the näive baseline is easier for certain locations, age groups, and time periods than others. To handle this difficulty, we use the *pairwise comparison* approach suggested in [17]. It is based on the assumption that achieving good nowcast performance *relative to all other considered methods* is similarly difficult across locations, age groups, and time periods. Considering a set of *N* models (including the baseline model), the relative WIS corrected for missing submissions is determined as follows:

1. In the first step for each pair of models *i, j* we compute the ratio

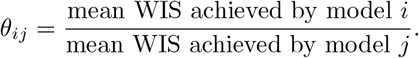
2. For each model *i* we then compute the geometric average of the ratios *θ*_*ij*_ achieved in the comparisons to all other models

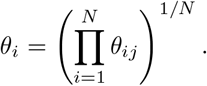
3. Lastly, we re-scale the *θ*_*i*_ to the one achieved by the baseline model to obtain the relative WIS:

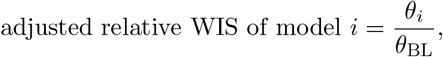

where *θ*_BL_ refers to the baseline model (`FrozenBaseline`).

If all models submitted all required nowcasts it is straightforward to show that this boils down to the regular relative WIS as defined in Section 2.5. If some submissions are missing for certain models, the procedure will adjust the relative WIS to how well other models fared on the respective subset of addressed targets.

#### c.2 Results

Table 6 compares the relative WIS computed using fill-in nowcasts as in the main analysis and the pairwise comparison approach. The differences are very modest, meaning that the missingness of nowcasts does not substantially affect the results. This is not surprising, given the low number of missing submissions.

### D Repositories of participating teams

`Epiforecasts`: https://epiforecasts.io/eval-germany-sp-nowcasting/
`KIT`: https://github.com/KITmetricslab/hospitalization-nowcast-hub/tree/main/code/baseline
`LMU`: https://github.com/MaxWeigert/Nowcasting_covid19_hospitalizations
`RIVM`: https://github.com/kassteele/Nowcast-hub
`SU`: https://github.com/FelixGuenther/hospitalization-nowcast-hub_SU-public

### E Documentation of the KIT model

As the `KIT` model was conceived as a conceptually simple (though not näive) reference model for the current study we provide a brief documentation of its methodology.

**Notation** Denote by *X*_*t,d*_, *d* = 0, …, *D* the number of hospitalizations for reference date *t* which appear in the data set at day *t* + *d* and by

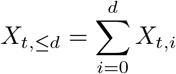

the number of hospitalizations reported for reference date *t* up to day *t* + *d*. Moreover, denote by

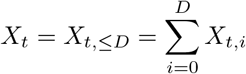

the total number of reported hospitalizations for *t*, where *D* denotes an assumed maximum possible delay. In the following, we denote by *X*_*t*_, etc. a random variable and by *x*_*t*_ the corresponding observation.

The observed *x*_*t,d*_ as available at a given time point *t*^*∗*^ can be arranged into the so-called *reporting triangle*, see Table 7.

**Table 7:** Illustration of the reporting triangle for time *t*^*∗*^ and *D* = 5. Quantities known at time *t* are shown in black, yet unknown quantities are shown in grey.

| day | $d = 0$ | $d = 1$ | $d = 2$ | $d = 3$ | $d = 4$ | $d = 5$ | total |
| --- | --- | --- | --- | --- | --- | --- | --- |
| 1 | $x_{1,0}$ | $x_{1,1}$ | $x_{1,2}$ | $x_{1,3}$ | $x_{1,4}$ | $x_{1,5}$ | $x_1$ |
| 2 | $x_{2,0}$ | $x_{2,1}$ | $x_{2,2}$ | $x_{2,3}$ | $x_{2,4}$ | $x_{2,5}$ | $x_2$ |
| ⋮ | | | | | | | |
| $t^* - 5$ | $x_{t^*-5,0}$ | $x_{t^*-5,1}$ | $x_{t^*-5,2}$ | $x_{t^*-5,3}$ | $x_{t^*-5,4}$ | $x_{t^*-5,5}$ | $x_{t^*-5}$ |
| $t^* - 4$ | $x_{t^*-4,0}$ | $x_{t^*-4,1}$ | $x_{t^*-4,2}$ | $x_{t^*-4,3}$ | $x_{t^*-4,4}$ | $x_{t^*-4,5}$ | $x_{t^*-4}$ |
| $t^* - 3$ | $x_{t^*-3,0}$ | $x_{t^*-3,1}$ | $x_{t^*-3,2}$ | $x_{t^*-3,3}$ | $x_{t^*-3,4}$ | $x_{t^*-3,5}$ | $x_{t^*-3}$ |
| $t^* - 2$ | $x_{t^*-2,0}$ | $x_{t^*-2,1}$ | $x_{t^*-2,2}$ | $x_{t^*-2,3}$ | $x_{t^*-2,4}$ | $x_{t^*-2,5}$ | $x_{t^*-2}$ |
| $t^* - 1$ | $x_{t^*-1,0}$ | $x_{t^*-1,1}$ | $x_{t^*-1,2}$ | $x_{t^*-1,3}$ | $x_{t^*-1,4}$ | $x_{t^*-1,5}$ | $x_{t^*-1}$ |
| $t^*$ | $x_{t^*,0}$ | $x_{t^*,1}$ | $x_{t^*,2}$ | $x_{t^*,3}$ | $x_{t^*,4}$ | $x_{t^*,5}$ | $x_{t^*}$ |

As we will focus on seven-day hospitalization incidences we moreover need to consider rolling sums over windows of length *W* (usually *W* = 7)

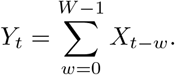

**Goal** Our aim is to estimate or *nowcast Y*_*t*_ based on the information available at time *t*^*∗*^ ≥ *t*. We do not take into account any information other than data on hospitalizations and their reporting delays, meaning that we model

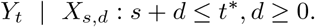

**Point nowcast** The following describes a simple heuristic to obtain a point prediction of *Y*_*t*_ based on information available at time *t*^*∗*^.

We start by imputing

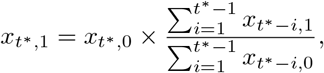

i.e. use a simple multiplication factor computed from the complete rows of our data set. Next, we compute

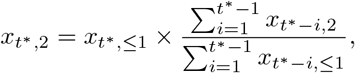

where in the computation of

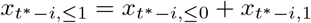

we just treat the *x*_*t∗−i*,1_ imputed in the first step as if it was a known value. The same can be done for

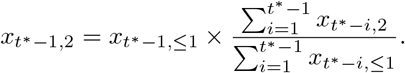

We repeat this same procedure to fill in the missing values of the reporting triangle step by step, moving from the left to the right and the bottom to the top.

This is equivalent to the following slightly more formal formulation: We denote by *π*_*d*_ the probability that a hospitalization with reference date *t* appears in the data on day *t* + *d* and by

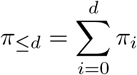

the probability that such a hospitalization appears in the data no later than *t* + *d*. We introduce

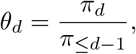

which allows us to formulate the recursion

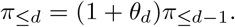

To estimate the *θ*_*d*_ for *d* = 1, …, *D < t* based on quantities available at time *t*^*∗*^ we use

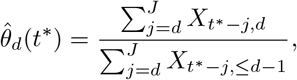

where *J* is the number of past observations to include in the estimation (in practice it is often helpful to use only a recent subset rather than the entire available history). Note that we treat this estimate as a function of *t*^*∗*^ as it may change over time. Estimates of the probabilities *π*_*≤d*_ can then be obtained as

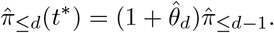

These can subsequently serve to estimate the total number *X*_*t*_ of hospitalizations with reference date *t* based on the *X*_*t,≤t∗−t*_ hospitalizations already reported by time *t*^*∗*^:

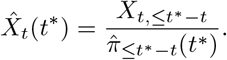

We can also compute the estimates for the respective number of hospitalizations reported with a given delay *d > t*^*∗*^ − *t*, which is given

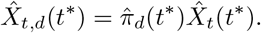

In the last step we move to the rolling sum *Y*_*t*_, which we estimate as

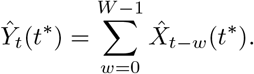

#### Uncertainty quantification

Our general idea to quantify the nowcast uncertainty for *Ŷ*_*t*_(*t*^*∗*^) is to generate point nowcasts *Ŷ*_*t−*1_(*t*^*∗*^ − 1), *Ŷ* _*t−*2_(*t*^*∗*^ − 2), …, *Ŷ* _*t−K*_ (*t*^*∗*^ − *K*) for *K > D* past time points, each based on the information available at the respective time point. These could then be compared to the correspond-ing observations *Y*_*t∗−*1_, …, *Y*_*t∗−K*_, and nowcast dispersion could be based on a simple parametric model. However, two aspects need to be taken into account:

- The information available at *t*^*∗*^, on which the nowcast *Ŷ*_*t*_(*t*^*∗*^) is based, already implies a lower bound for *Y*_*t*_, namely the hospitalizations which have already been observed. Only the hospitalizations for reference date *t* which will be reported after *t*^*∗*^ need to be modeled probabilistically. We thus introduce the decomposition

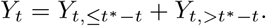

Here,

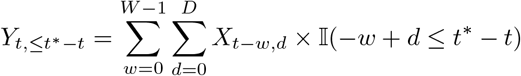

are those already observed by *t*^*∗*^ (i.e., the lower bound) and

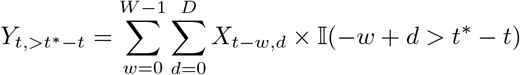

are those yet to be observed. We only need to quantify the uncertainty about the latter.
- At time *t*^*∗*^, the realizations of *Y*_*t,>t∗−t*_ are only available for *t* ≤ *t*^*∗*^ − *D*. If we only want to use complete observations we would need to discard a lot of recent information. We therefore construct a set of observations *Z*_*t−j,>t∗−t*_, *j* = 1, …, *K* and corresponding point predictions 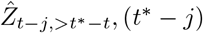 as follows:
- For *j* = *D*, …, *K* we can simply set

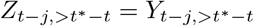

and point predictions 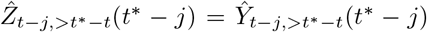 as all relevant information are already available at *t*^*∗*^.
- For *j* = 1, …, *D* – 1 we use partial observations

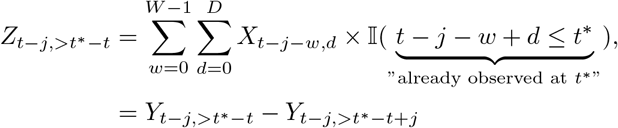

which are restricted to hospitalizations already reported by time *t*^*∗*^, so that *Z*_*t−j,>t∗−t*_ can be evaluated. The corresponding point nowcasts are given by

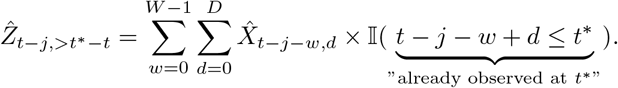

We then pragmatically assume that

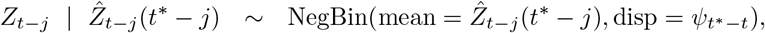

where we parameterize the negative binomial distribution via its mean and the dispersion (size) parameter *?*_*t∗−t*_. Note that the dispersion parameter depends on how far back into the past we nowcast (i.e., how much information has already accumulated between *t* − *j* and *t*^*∗*^ − *j*). The parameters *?*_0_, …, *?*_*D*_ are then estimated via maximum likelihood. To avoid issues with zero expectations we add 0.1 to the expected values when feeding them into the maximum likelihood procedure.

The predictive distributions for *Y*_*t*_ are then set to NegBin(mean = *Ŷ*_*t,>t∗ −t*_(*t*^*∗*^), size = *?*_*t∗−t*_), shifted by *Y*_*t,≤t∗−t*_. As a motivation for the use of partial observations in the estimation of the overdispersion parameters, we note that if

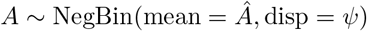

And

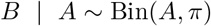

one gets

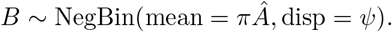

The negative binomial distribution with a given dispersion parameter is thus closed to binomial subsampling, with only the expectation, but not the size parameter changing. It is therefore defendable to assume the same size parameter for the constructed partial observations *Z*_*t−j,>t∗−t*_ and the actual *Y*_*t−j,>t∗−t*_ which we would use if they were already available.

#### Parameter choices

To apply the suggested method, the numbers *J* and *K* of past observations are used to estimate the nowcast mean and dispersion parameters. Here one needs to strike a balance between a sufficient amount and recency of training data. We set both *J* and *K* to 60 days without further assessing the impact on nowcast quality. The maximum delay of *D* was set to 40 days.

## References

[1] D. Giannone, L. Reichlin, and D. Small, “Nowcasting: The real-time informational content of macroe-conomic data,” Journal of Monetary Economics, vol. 55, no. 4, pp. 665–676, 2008.

[2] P. D. England and R. J. Verrall, “Stochastic claims reserving in general insurance,” British Actuarial Journal, vol. 8, no. 3, pp. 443–518, 2002.

[3] D. R. Cox and G. F. Medley, “A process of events with notification delay and the forecasting of AIDS,” Philosophical Transactions of the Royal Society of London. B, Biological Sciences, vol. 325, no. 1226, pp. 135–145, 1989.

[4] M. Höhle and M. an der Heiden, “Bayesian nowcasting during the STEC O104:H4 outbreak in Germany, 2011,” Biometrics, vol. 70, no. 4, pp. 993–1002, 2014.

[5] T. Donker, M. van Boven, W. van Ballegooijen, T. van’t Klooster, et al., “Nowcasting pandemic influenza A/H1N1 2009 hospitalizations in the Netherlands,” European Journal of Epidemiology, vol. 26, no. 3, pp. 195–201, 2011.

[6] T. F. Menkir, H. Cox, C. Poirier, M. Saul, et al., “A nowcasting framework for correcting for reporting delays in malaria surveillance,” PLoS Computational Biology, vol. 17, no. 11, e1009570, 2021.

[7] L. S. Bastos, T. Economou, M. F. Gomes, D. A. Villela, et al., “A modelling approach for correcting reporting delays in disease surveillance data,” Statistics in Medicine, vol. 38, no. 22, pp. 4363–4377, 2019.

[8] S. F. McGough, M. A. Johansson, M. Lipsitch, and N. A. Menzies, “Nowcasting by Bayesian smoothing: A flexible, generalizable model for real-time epidemic tracking,” PLoS computational biology, vol. 16, no. 4, e1007735, 2020.

[9] F. Gunther, A. Bender, K. Katz, H. Kuchenhoff, and M. Höhle, “Nowcasting the COVID-19 pandemic in Bavaria,” Biometrical Journal, vol. 63, no. 3, pp. 490–502, 2021.

[10] S. R. Seaman, P. Samartsidis, M. Kall, and D. De Angelis, “Nowcasting COVID-19 deaths in England by age and region,” Journal of the Royal Statistical Society: Series C (Applied Statistics), vol. 71, no. 5, pp. 1266–1281, 2022.

[11] R. Jersakova, J. Lomax, J. Hetherington, B. Lehmann, et al., “Bayesian imputation of COVID-19 positive test counts for nowcasting under reporting lag,” Journal of the Royal Statistical Society: Series C (Applied Statistics), vol. 71, no. 4, pp. 834–860, 2022.

[12] T. Li and L. F. White, “Bayesian back-calculation and nowcasting for line list data during the COVID-19 pandemic,” PLOS Computational Biology, vol. 17, no. 7, pp. 1–22, Jul. 2021.

[13] I. Hawryluk, H. Hoeltgebaum, S. Mishra, X. Miscouridou, et al., “Gaussian process nowcasting: Application to COVID-19 mortality reporting,” in Proceedings of the Thirty-Seventh Conference on Uncer-tainty in Artificial Intelligence, C. de Campos and M. H. Maathuis, Eds., ser. Proceedings of Machine Learning Research, vol. 161, PMLR, 27–30 Jul 2021, pp. 1258–1268.

[14] S. K. Greene, S. F. McGough, G. M. Culp, L. E. Graf, et al., “Nowcasting for real-time COVID-19 tracking in New York City: An evaluation using reportable disease data from early in the pandemic,” JMIR Public Health Surveill, vol. 7, no. 1, e25538, Jan. 2021, issn: 2369-2960.

[15] German Federal Government, “Videoschaltkonferenz der Bundeskanzlerin mit den Regierungschefinnenund Regierungschefs der Lander am 18. November 2021,” Tech. Rep., 2021,https://www.bundesregierung.de/resource/blob/974430/1982598/defbdff47daf5f177586a5d34e8677e8/2021-11-18-mpk-data.pdf.

[16] M. Milatz and I. Lerch, “Nach MPK-Beschluss: Verwirrung um Hospitalisierungsinzidenz,” Nord-deutscher Rundfunk, Tech. Rep., 2021, https://www.ndr.de/nachrichten/info/Nach-MPK-Beschluss-Verwirrung-um-Hospitalisierungsinzidenz,hospitalisierungsinzidenz100.html.

[17] E. Y. Cramer, E. L. Ray, V. K. Lopez, J. Bracher, et al., “Evaluation of individual and ensemble probabilistic forecasts of COVID-19 mortality in the United States,” Proceedings of the National Academy of Sciences, vol. 119, no. 15, e2113561119, 2022.

[18] J. Bracher, D. Wolffram, J. Deuschel, K. Görgen, et al., “A pre-registered short-term forecasting study of COVID-19 in Germany and Poland during the second wave,” Nature Communications, vol. 12, no. 1, p. 5173, 2021.

[19] K. Sherratt, H. Gruson, H. Johnson, R. Niehus, et al., “Predictive performance of multi-model ensemble forecasts of COVID-19 across European nations,” medRxiv, 2022, doi:https://doi.org/10.1101/ 2022.06.16.22276024s.

[20] N. G. Reich, L. C. Brooks, S. J. Fox, S. Kandula, et al., “A collaborative multiyear, multimodel assessment of seasonal influenza forecasting in the United States,” Proceedings of the National Academy of Sciences, vol. 116, no. 8, pp. 3146–3154, 2019.

[21] J. Bracher, E. L. Ray, T. Gneiting, and N. G. Reich, “Evaluating epidemic forecasts in an interval format,” PLOS Computational Biology, vol. 17, no. 2, e1008618, 2021.

[22] J. Bracher, D. Wolffram, the COVID-19 Nowcast Hub Team, and Participants, Study protocol: Comparison and combination of COVID-19 hospitalization nowcasts in Germany, Deposited 23 November 2021, Registry of the Open Science Foundation, https://osf.io/mru75/, 2021.

[23] Robert Koch Institute, COVID-19-Hospitalisierungen in Deutschland, available online at https://github.com/robert-koch-institut/COVID-19-Hospitalisierungen_in_Deutschland, DOIhttps://doi.org/10.5281/zenodo.7527802, 2022.

[24] German Federal Ministry of Health, “FAQ zur Hospitalisierungsinzidenz,” Tech. Rep., 2021,https://www.bundesgesundheitsministerium.de/coronavirus/hospitalisierungsinzidenz.html.

[25] K. Tolksdorf, W. Haas, E. Schuler, L. H. Wieler, et al., “Syndromic surveillance for severe acute respiratory infections (SARI) enables valid estimation of COVID-19 hospitalization incidence and reveals underreporting of hospitalizations during pandemic peaks of three COVID-19 waves in Germany, 2020-2021,” medRxiv, 2022.

[26] S. Abbott, A. Lison, and S. Funk, “Epinowcast: Flexible hierarchical nowcasting,” Zenodo, 2021. doi:10.5281/zenodo.5637165.

[27] C. Fritz, G. D. Nicola, M. Rave, M. Weigert, et al., “Statistical modelling of COVID-19 data: Putting generalized additive models to work,” Statistical Modelling, no. forthcoming, 2023.

[28] M. Schneble, G. De Nicola, G. Kauermann, and U. Berger, “Nowcasting fatal COVID-19 infections on a regional level in Germany,” Biometrical Journal, vol. 63, no. 3, pp. 471–489, 2021.

[29] J. van de Kassteele, P. H. Eilers, and J. Wallinga, “Nowcasting the number of new symptomatic cases during infectious disease outbreaks using constrained p-spline smoothing,” Epidemiology (Cambridge, Mass.), vol. 30, no. 5, p. 737, 2019.

[30] M. an der Heiden and O. Hamouda, “Schatzung der aktuellen Entwicklung der SARS-CoV-2-Epidemie in Deutschland – Nowcasting,” Epidemiologisches Bulletin, vol. 2020, no. 17, pp. 10–15, 2020. doi: http://dx.doi.org/10.25646/6692.4

[31] J. Lawless, “Adjustments for reporting delays and the prediction of occurred but not reported events,” Canadian Journal of Statistics, vol. 22, no. 1, pp. 15–31, 1994.

[32] C. Genest, “Vincentization revisited,” The Annals of Statistics, pp. 1137–1142, 1992.

[33] K. C. Lichtendahl, Y. Grushka-Cockayne, and R. L. Winkler, “Is it better to average probabilities or quantiles?” Management Science, vol. 59, no. 7, pp. 1594–1611, 2013.

[34] T. Gneiting and A. E. Raftery, “Strictly proper scoring rules, prediction, and estimation,” Journal of the American Statistical Association, vol. 102, no. 477, pp. 359–378, 2007.

[35] W. Ehm, T. Gneiting, A. Jordan, and F. Kruger, “Of quantiles and expectiles: Consistent scoring functions, Choquet representations and forecast rankings,” Journal of the Royal Statistical Society: Series B (Statistical Methodology), vol. 78, no. 3, pp. 505–562, 2016.

[36] G. Shafer and V. Vovk, “A tutorial on conformal prediction,” Journal of Machine Learning Research, vol. 9, no. 12, pp. 371–421, 2008.

[37] Berliner Morgenpost, “Triage in Sachsen: Kliniken bereiten sich auf Schlimmes vor,” Berliner Morgenpost, 2021, published online on 23 November 2021, https://www.morgenpost.de/vermischtes/ article233915811/corona-sachsen-triage-intensivstationen-ueberlastung.html.

[38] N. Wolter, W. Jassat, S. Walaza, R. Welch, et al., “Early assessment of the clinical severity of the SARS-CoV-2 omicron variant in South Africa: A data linkage study,” The Lancet, vol. 10323, pp. 437–446, 2022.

[39] F. Bergström, F. Gunther, M. Höhle, and T. Britton, “Bayesian nowcasting with leading indicators applied to COVID-19 fatalities in Sweden,” PLOS Computational Biology, vol. 18, no. 12, pp. 1–17, Dec. 2022.

[40] M. Heinsch and J. Schmid-Johannsen, Mit oder wegen Corona im Krankenhaus? So bedingt aussagekraftig sind die BW-Daten, available online, https://www.swr.de/swraktuell/baden-wuerttemberg/was-sagt-die-hospitalisierungsinzidenz-in-der-omikron-welle-aus-100.html, 4 February 2022, 2022.

[41] S. Abbott, A. Lison, S. Funk, C. Pearson, and H. Gruson, “Epinowcast: Flexible hierarchical nowcasting,” Zenodo, 2021. doi: 10.5281/zenodo.5637165. [Online]. Available: https://github.com/epinowcast/epinowcast.

